# Associations Between TMS-Induced Electric Fields and Craving and Consumption Outcomes in Substance Use Disorders: A Multimodal Dose-Response Meta-Analysis

**DOI:** 10.64898/2026.06.23.26356355

**Authors:** Ghazaleh Soleimani, Martin P Paulus, Hamed Ekhtiari, Alexander Opitz

## Abstract

**Background:** Transcranial magnetic stimulation (TMS) is a promising treatment for substance use disorders (SUDs), although heterogeneous stimulation parameters hinder the identification of optimal strategies. Using meta-modeling, we linked treatment effect sizes (Hedges’ g) to simulated electric field (E-field) distributions to identify brain regions associated with efficacy variability.

**Methods:** TMS trials in individuals with SUDs published through the end of 2025 were identified through a systematic PubMed search. Studies reporting craving or consumption outcomes with quantifiable effect sizes were included. Objectives were to (i) examine associations between study-level effect sizes and simulated local E-field strength in MNI space for craving and consumption outcomes; (ii) generate a combined E-field–effect size association map; and (iii) assess spatial overlap with fMRI drug cue–reactivity patterns in 60 individuals with SUDs.

**Results:** The analysis included 81 randomized controlled TMS studies, yielding 107 effect size estimates for craving and consumption (n = 75 and n = 32, respectively). Compared with sham stimulation, TMS produced small-to-moderate improvements in both outcomes. E-field modeling identified the pre-supplementary motor area (pre-SMA) and inferior frontal gyrus (IFG) as regions associated with variability in craving-related effect sizes, and the frontopolar cortex with variability in consumption-related effect sizes. Correlation maps were highly robust (mean leave-one-out similarity r = 0.996), and the frontopolar cluster showed significant spatial overlap with fMRI drug cue–reactivity patterns (Dice coefficient = 0.37).

**Conclusion:** These findings identify frontopolar, pre-SMA, and IFG regions where local E-field strength is associated with SUD treatment effects, supporting more precise neuromodulation strategies.

## Introduction

Substance use disorders (SUDs) represent a persistent and widespread global public health burden with substantial individual and societal impact. Growing insights into the neurobiology of addiction have supported the development of pharmacological interventions targeting specific neurotransmitter systems implicated in SUDs(1). Complementing these advances, non-invasive brain stimulation (NIBS) approaches, most notably transcranial magnetic stimulation (TMS), have gained increasing attention as potential adjunctive treatments(2). By generating rapidly changing magnetic fields, TMS induces electric fields and secondary currents within cortical tissue through electromagnetic induction, thereby influencing neural activity within circuits implicated in cue reactivity and addictive behaviors (3, 4, 5). The spatial extent of the induced electric field depends on coil geometry, stimulation parameters, and individual anatomy. Figure-of-eight coils generally produce more spatially focused electric fields than circular or deep-TMS coils, although stimulation remains distributed across several centimeters of cortical tissue (6, 7). Previous meta-analyses have reported small-to-moderate effect sizes for TMS in reducing craving and substance consumption(8, 9). However, substantial inter-experimental variability limits the generalizability of findings and their translation into optimized stimulation protocols.

Given the large parameter space associated with TMS interventions, this inter-experimental variability may arise from multiple sources, including differences in stimulation parameters, including the targeted brain region, stimulation intensity, coil type (10, 11), coil orientation(12), and stimulation protocol (e.g., high frequency, low frequency, or theta burst stimulation)(13), as well as variation in targeting methods, cumulative stimulation dose, treatment duration, participant characteristics, and concurrent treatments. The spatial distribution of the TMS-induced electric field (E-field) and, consequently, the brain circuits that are engaged can vary substantially in response to relatively small variations in these parameters. Furthermore, the neurophysiological effects of different TMS protocols exhibit substantial inter-individual and state-dependent variability, such that their net effects cannot be reliably inferred solely from stimulation frequency or burst structure (see Supplementary Section S1 for examples).

To address this limitation, we implemented a meta-modeling framework designed to estimate the association between study-level treatment effect sizes (Hedges’ g) and study-level simulated electric field (E-field) distributions (Figure 1). We conducted a systematic review and meta-analysis of TMS studies in SUDs reporting craving or consumption outcomes, yielding 107 effect size estimates. For each study, TMS-induced E-field distributions were simulated using study-specific stimulation parameters, including target location, coil type, coil orientation, stimulation intensity, and dI/dt (the rate of change of current flowing through the stimulation coil, which determines the induced electric field strength), in a standard head model (Ernie) and a clinically representative cohort of 60 individuals with methamphetamine use disorder, and subsequently transformed into MNI space for group-level analyses. A previously validated correlation-based approach(14, 15, 16, 17) was then used to relate between-study variability in local E-field strength to variability in behavioral effect sizes across the brain.

**Figure 1.**
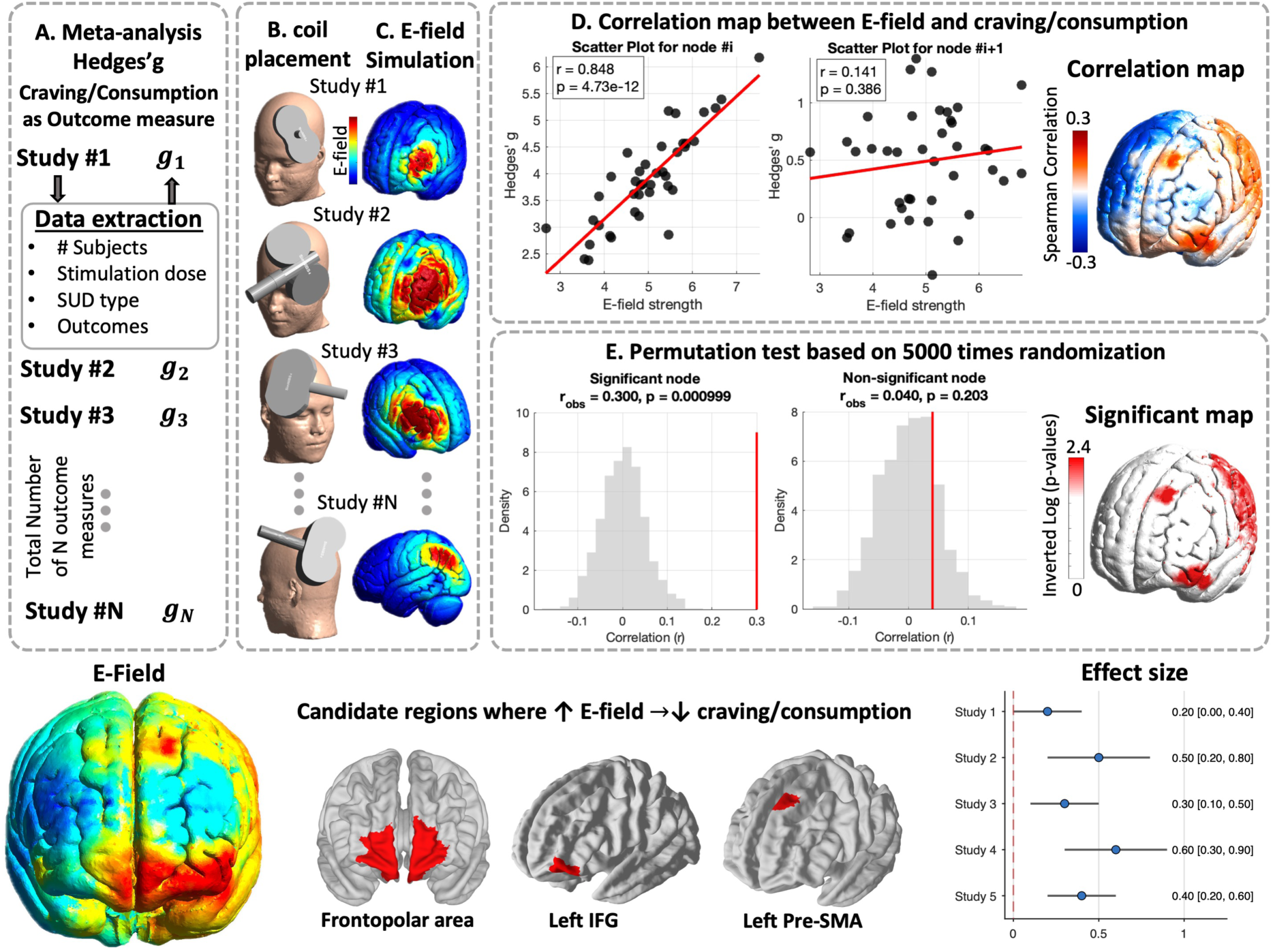
Overview of the meta-modeling pipeline for mapping E-field–behavior associations in TMS studies for substance use disorders (SUD). **(A) Standard meta-analysis step.** For each eligible randomized clinical trial, effect sizes (Hedges’ g) indexing changes in substance craving or consumption were extracted along with stimulation parameters, study characteristics, and outcomes. Each study contributed one effect size (g_1_…g_n_) to the dataset. **(B) Coil placement.** Stimulation targets and coil positions were reconstructed based on the targeting approach reported in each study. **(C) Computational modeling step.** Based on the reported coil type, stimulation target, coil orientation, stimulation intensity, and dI/dt (rate of change of coil current), TMS-induced electric-field (E-field) simulations were generated using anatomically realistic head models. Simulated E-field maps were transformed into a common space to enable comparisons across studies. **(D) Correlation mapping.** Across all studies, node-wise Spearman correlations were computed between local E-field strength and corresponding Hedges’ g values, generating a whole-brain map of E-field–outcome associations. Example scatter plots illustrate positive and weak associations at individual cortical nodes. **(E) Statistical significance testing.** A permutation-based framework was applied in which effect size labels were randomly shuffled 5,000 times relative to E-field maps to generate empirical null distributions. Observed correlations were compared against these null distributions to identify significant cortical regions. The resulting significant regions represent candidate locations where variability in E-field engagement was associated with variability in treatment outcomes across studies.

The <u>primary objective</u> of this study was to estimate these E-field–effect size associations separately for craving and consumption outcomes, given their distinct clinical and behavioral interpretation. <u>Secondary objectives</u> were twofold. (I) We generated a combined E-field–effect size association map by integrating craving and consumption outcomes to provide a descriptive summary across symptom domains. (II) We evaluated the spatial overlap between E-field–informed association maps and fMRI drug cue reactivity patterns in 60 individuals with methamphetamine use disorder (MUD) to assess external consistency. For this purpose, individualized head models (60×107 simulations) were constructed to assess spatial correspondence between E-field–derived association maps and task-related neural activity.

We hypothesized that variability in TMS-induced E-fields would be associated with variability in behavioral outcomes across studies and correspond to functionally relevant neural systems. To test this hypothesis, we applied an E-field–based meta-modeling framework linking study-level effect sizes to simulated E-field distributions. Given the observational nature of these analyses, findings should be considered hypothesis-generating rather than causal.

## Method

### Study Identification and Effect Size Extraction

Following our previous work (8), we conducted an updated systematic review of TMS studies in substance use disorders according to PRISMA guidelines(18). Eligible studies were identified through a systematic literature search, and stimulation parameters, study characteristics, and outcome measures were extracted (Table S1). For each study, behavioral outcomes were summarized as Hedges’ g using the *metafor* package in R. When multiple measures of the same outcome were reported, effect sizes were combined to obtain a single independent estimate per outcome. Additional details regarding search strategy, data extraction, inclusion/exclusion criteria, effect size calculations, risk-of-bias assessment, and statistical procedures are provided in Supplementary Sections S2–S4.

### Electric Field (E-field) Modeling

High-resolution T1-weighted MRI data and SimNIBS 4.1 software (19) were used to simulate the TMS-induced E-field in standard MNI space. In each study, the TMS-induced E-field strength was simulated based on considering the TMS target location, coil orientation, stimulation intensity, and coil type. In addition to the MNI-space analyses, this pipeline was replicated in a sample healthy subject (“Ernie”), available in the SimNIBS example dataset, to ensure the robustness and reproducibility of our results in an individual head model. Analyses were also repeated across 60 anatomically individualized head models derived from participants with MUD to assess inter-subject variability for a real SUD population. Details about head modeling and inter-individual variability considerations are provided in Supplementary Section S5. Additional details on the MRI data are provided in Supplementary Section S6, and further demographic information for the SUD population is available in Supplementary Section S7.

Across the dataset, 107 effect size estimates (59 craving-related and 48 consumption-related) were extracted and linked to stimulation targets spanning the prefrontal, parietal, and temporal cortices, implemented using diverse coil designs, orientations, and stimulation intensities. Because different TMS coils generate different levels of E-field strengths, E-field values were normalized before analysis; these normalized values were used both to generate average E-field maps and to compute E-field–outcome correlations. To ensure that this normalization does not affect the final correlation map, the pipeline was replicated without normalization (Supplementary Sections S13–S14).

### E-field–Effect Size Association Maps

The primary analysis focused on estimating associations between study-level treatment effect sizes (Hedges’ g) and study-level simulated local electric field (E-field) strength across cortical locations, while all additional analyses were considered secondary and hypothesis-generating.

All analyses were conducted at the study level, relating between-study variability in simulated E-field strength to variability in reported effect sizes. To achieve this, we implemented a node-wise correlation analysis following established and validated procedures(14, 15, 16, 17, 20). For each cortical node (triangular surface element), local E-field strength values were paired with study-specific effect size estimates (Hedges’ g), reflecting behavioral outcomes associated with the corresponding stimulation parameters.

Analyses were conducted in standard brain space using a cortical mesh comprising 986,936 surface nodes. Across the 107 effect size estimates, this resulted in a 986,936 × 107 E-field matrix per head model, where each column represented the spatial distribution of induced fields associated with a given experimental condition. Rank-based Spearman correlations were computed between local E-field strength and corresponding vectors of Hedges’ g values across studies. Analyses were performed on selected subsets where appropriate (e.g., studies employing specific coil types). Because individual-level participant data were not available, within-study covariance between multiple reported outcomes could not be estimated. Therefore, effect sizes were treated as independent observations, and this assumption was evaluated using a one-effect-per-study sensitivity analysis. Correlation coefficients quantify the association between local E-field strength and variability in behavioral effect sizes across studies. Positive correlation values indicate that higher local E-field strength is associated with larger reductions in craving or consumption (i.e., greater effect sizes), whereas negative values indicate associations with smaller or adverse effect sizes. For reporting purposes, node-wise correlation estimates were summarized across anatomical cortical regions and subregions.

### Primary Analysis: Outcome-Specific Maps

Outcome-specific analyses were conducted using craving- and consumption-related effect size estimates (Hedges’ g) as distinct primary outcomes, given their differences in clinical and behavioral interpretation. The node-wise correlation procedures described above were applied separately to the subsets of studies reporting craving and consumption outcomes to estimate associations between local E-field strength and variability in outcome-specific effect sizes across studies. Results were interpreted independently for each outcome domain.

### Secondary Analysis I: Combined E-field–Effect Size Association Map

As a secondary analysis, a pooled E-field–effect size association map was generated by including all eligible effect size estimates derived from both craving and consumption outcomes. Specifically, rather than restricting the analysis to a single outcome domain, all effect size estimates were pooled into a single dataset. The same node-wise correlation framework described above was applied to the pooled set of effect sizes to estimate associations between local E-field strength and variability in combined behavioral outcomes across studies. This analysis was conducted for descriptive comparison across symptom domains and was not intended to reflect a unified biological mechanism underlying craving and consumption. Instead, it provides a summary representation of spatial patterns of association across outcome types.

### Secondary Analysis II: Spatial Overlap with fMRI Drug Cue Reactivity

As a secondary analysis, fMRI data were used to assess spatial correspondence between E-field–effect size association maps and cue-reactivity–related neural activity. For the 60 participants with methamphetamine use disorder (MUD) whose T1-weighted anatomical scans were used for individualized head-model construction, fMRI data were obtained using a standardized drug cue reactivity (FDCR) paradigm. During the task, participants viewed pictorial stimuli depicting methamphetamine-related cues and neutral control images arranged in a block design consisting of four blocks per condition. Each block contained six images, each presented for 5 seconds with short interstimulus intervals, separated by fixation periods of 8–12 seconds; the total task duration was approximately 6.5 minutes. Task regressors were constructed by convolving block timing with a boxcar function. A supplementary model incorporating additional trial- and rating-phase regressors yielded consistent results. First-level general linear model (GLM) analyses were performed, and group-level FDCR maps were derived using the drug > neutral contrast. For visualization and comparison with E-field association maps, the resulting activation maps were thresholded to display beta coefficients within the range of −3 to 3. Additional details are provided in Supplementary Section S8. This analysis was intended to assess spatial correspondence and should not be interpreted as validation of causal targets or individualized treatment efficacy.

To quantify spatial correspondence between FDCR activation maps and E-field–behavior association maps, surface-based correlation clusters obtained from the 60 × 107 E-field analyses were converted to volumetric space using the “msh2nii” function. Clusters derived from the correlation analyses were projected into volume space and binarized separately for craving- and consumption-related effects. Similarly, group-level FDCR activation maps were thresholded to identify regions showing cue-reactivity–related activation. Because the goal of this analysis was to evaluate the spatial overlap between regions identified by the two approaches, rather than to compare continuous-valued E-field association strengths and fMRI activation amplitudes on a voxel-wise basis, both maps were represented as binary spatial masks. Spatial overlap between the resulting E-field association masks and FDCR activation masks was quantified using the Dice similarity coefficient, a measure commonly used to assess overlap between thresholded spatial regions. This analysis was designed to determine whether regions identified by the E-field–effect size associations overlapped more than expected by chance with regions exhibiting cue-reactivity–related activation, and should not be interpreted as a measure of voxel-wise correspondence between continuous maps. Because random voxel-wise shuffling does not preserve the spatial autocorrelation structure of fMRI data, Dice coefficients were interpreted descriptively rather than as formal inferential statistics. To assess whether spatial overlap was sensitive to threshold choice, overlap analyses were repeated across multiple percentile-based thresholds.

### Statistical Inference and Robustness Analyses

Statistical significance of E-field–effect size associations was evaluated using permutation testing with correction for multiple comparisons across cortical nodes. Robustness was assessed using leave-one-out and K-fold cross-validation procedures, sensitivity analyses addressing within-study dependence, and threshold-dependent spatial overlap analyses. Full methodological details about statistical analysis are provided in Supplementary Section S9.

## Results

The PRISMA flowchart detailing study selection, rejection, and key variables is shown in Figure 2. None of the studies were classified as ’high risk,’ and only two were marked as having ’some concerns.’ The final dataset included 107 effect size estimates from 81 randomized and controlled TMS trials in substance use disorders.

**Figure 2.**
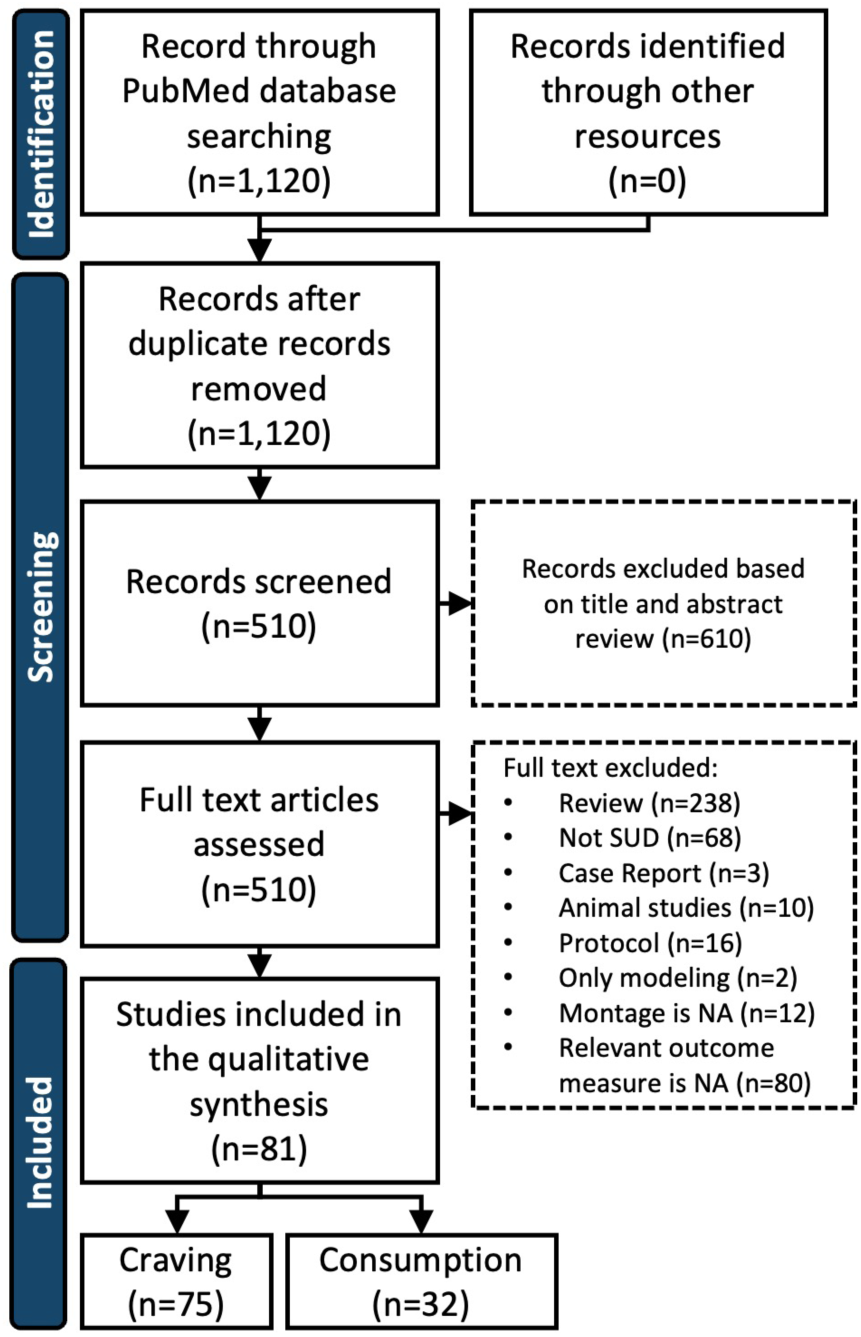
PRISMA flow diagram. Summarizing the selection of all published studies applying transcranial magnetic stimulation (TMS) for substance use disorders (SUDs) with reported craving or consumption outcomes. *n* is the number of studies; however, in the craving and consumption report, *n* is the number of included outcome measures.

### Study Characteristics

Studied substances were predominantly nicotine (41.1%, n = 44) and alcohol (22.4%, n = 24), followed by amphetamines, opioids, cocaine, and cannabis. Stimulation protocols varied substantially across studies, with sessions ranging from 1 to 30 (median = 10) and frequencies between 1 and 20 Hz (median = 10 Hz). Conventional rTMS was the most common modality (57.9%, n = 62), followed by theta-burst stimulation (28.0%, n = 30) and deep TMS (14.0%, n = 15).

Outcomes were primarily assessed immediately post-stimulation (95.3%), with relatively few delayed follow-ups, indicating that the literature predominantly reflects acute effects. Most effect sizes corresponded to craving outcomes (70%, n = 75), with fewer assessing consumption (30%, n = 32). Outcome measures were heterogeneous and included visual analogue scales (VAS), self-reported substance use, and validated craving instruments. More details can be found in Supplementary Sections S10 and S11.

### Electric Field Distribution Across Studies

Figure 3 illustrates the spatial distribution of mean normalized E-field strength across stimulation conditions on a standard brain template. For conventional coils, peak cortical E-field strength (99.9th percentile) reached 136.4 V/m in the left dorsolateral prefrontal cortex (MNI −42.3, 43.6, 27.4). For deep TMS, peak cortical E-field strength reached 155.8 V/m in the right frontal cortex (MNI 28.7, 41.4, 28.9). See Supplementary Section S12 for details of E-field modeling and stimulation parameters, Supplementary Section S13 for mean and variability of E-field distributions across commonly targeted brain regions, and Supplementary Section S14 for differences in E-field distributions between conventional and deep TMS coils.

**Figure 3.**
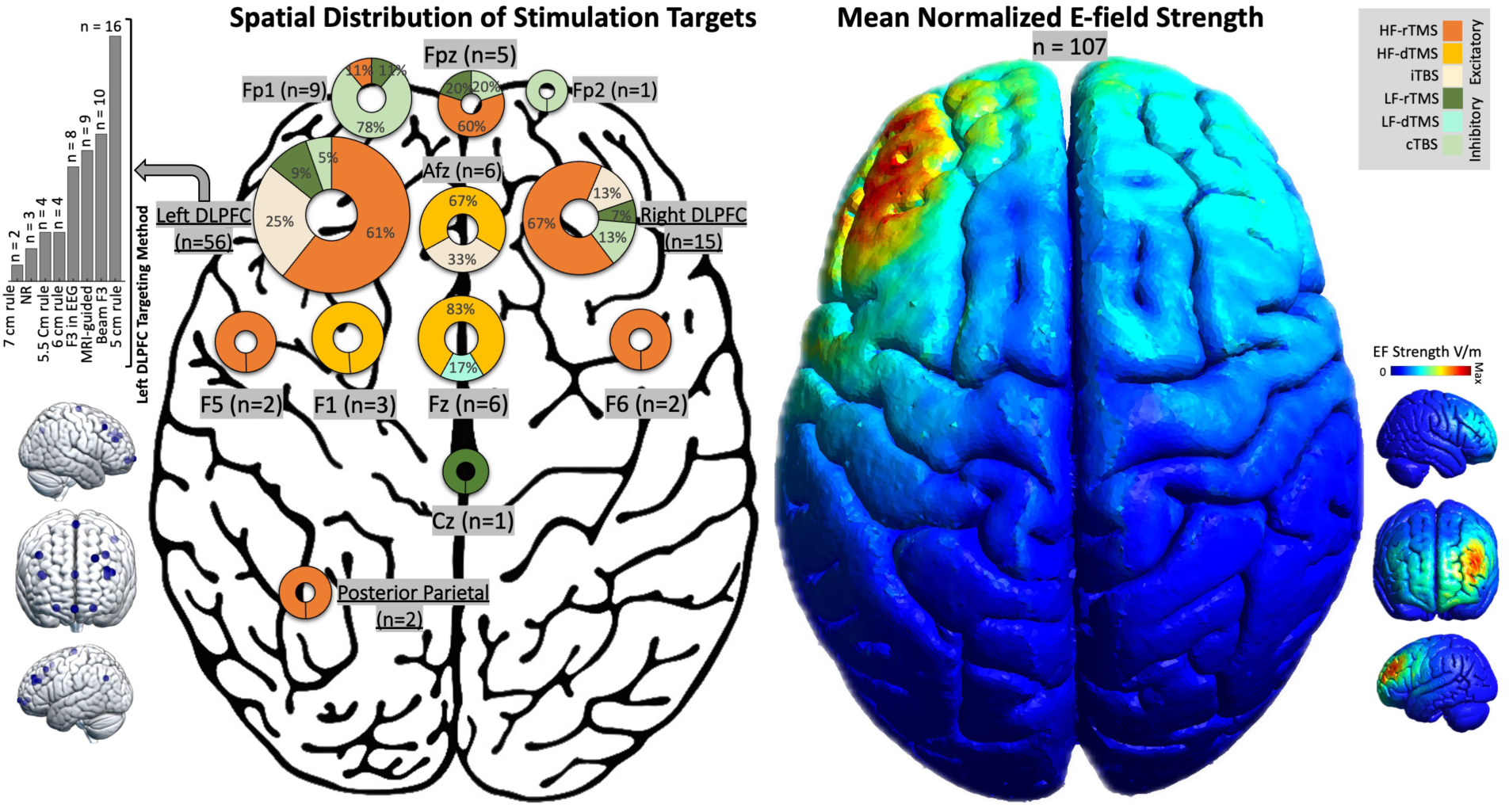
Spatial distribution of stimulation targets and cortical patterns of normalized electric field strength. **<u>Left panel:</u>** Schematic illustration of the spatial distribution of stimulation targets projected onto a standardized cortical surface. Circles represent commonly targeted scalp or cortical locations, positioned according to their anatomical correspondence. The size of each circle reflects the relative prominence of that target location across studies, while color shading within the circles encodes stimulation characteristics. Colors indicate different stimulation protocol categories. Warm colors (yellow–orange) represent high-frequency stimulation protocols (e.g., HF-rTMS and iTBS), whereas cooler or muted tones (green–olive) represent low-frequency stimulation protocols (e.g., LF-rTMS and cTBS). Protocols are categorized according to stimulation pattern rather than presumed neurophysiological effects. Small brain icons adjacent to the schematic provide spatial context for cortical orientation and hemispheric representation. The bar plot summarizes the relative use of different stimulation targeting approaches reported across the literature. Each bar represents a distinct targeting method, grouped by conceptual similarity (e.g., scalp-based heuristics versus individualized approaches). Bar height reflects the relative prevalence of each targeting strategy. **<u>Right panel:</u>** Three-dimensional cortical surface rendering illustrating the mean normalized electric field (E-field) strength across stimulation conditions, displayed on a standard brain template. The color map shows the spatial distribution of the normalized E-field magnitude, with warm colors (yellow to red) indicating regions of relatively higher induced E-field strength and cool colors (light to dark blue) indicating regions of lower E-field strength. Consistent with current evidence, the neurophysiological effects of TMS protocols may vary substantially across individuals, brain states, cortical targets, and stimulation parameters; therefore, physiological effects cannot be reliably inferred solely from stimulation frequency or burst structure.

**Figure 4.**
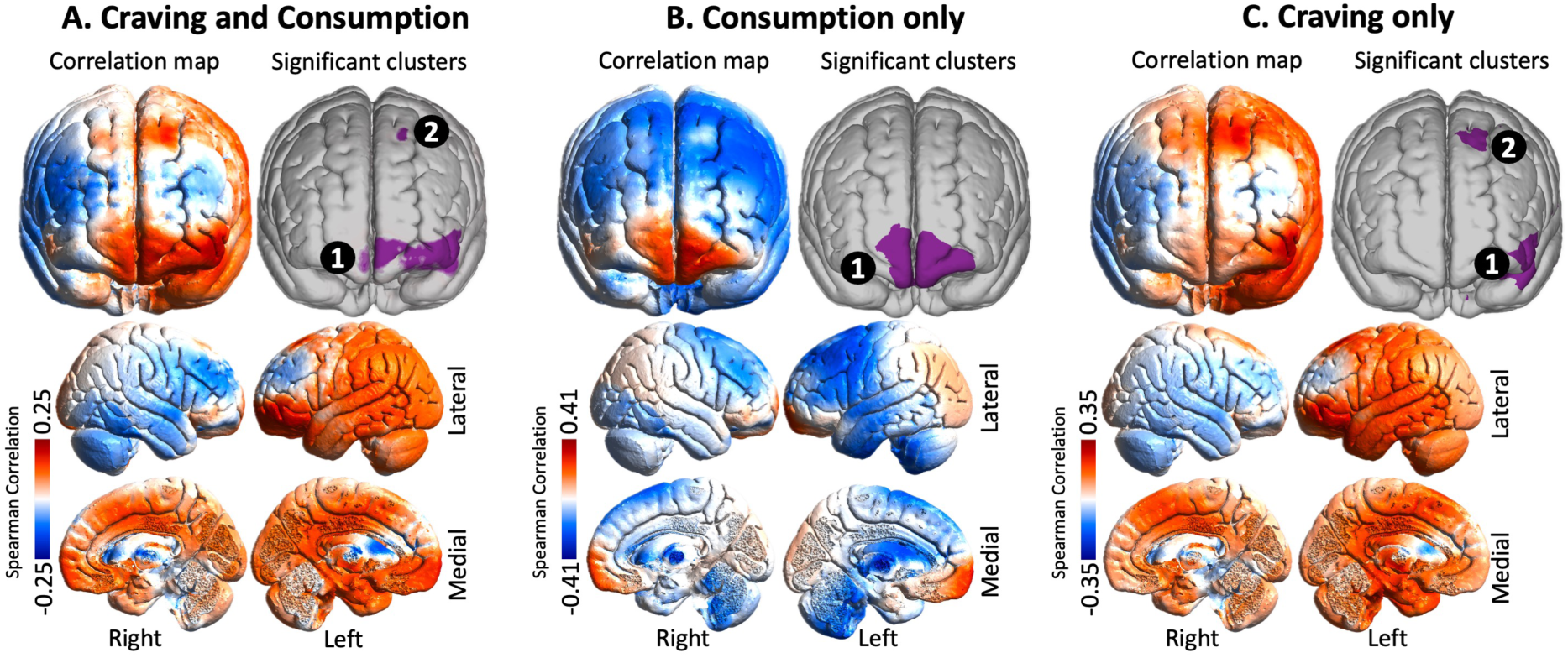
Electric field–outcome correlation maps for transcranial magnetic stimulation across substance use disorder studies. Whole-brain Spearman correlation maps between simulated cortical electric field (E-field) strength and clinical effect sizes (Hedges’ g) were computed across studies at each cortical node using anatomically normalized E-field maps. Warmer colors indicate positive correlations (higher E-field strength associated with larger clinical benefit), whereas cooler colors indicate negative correlations. Purple overlays denote cortical clusters surviving permutation-based multiple-comparison correction. **(A) Craving only.** Correlation maps and significant clusters derived from studies reporting craving outcomes only. Significant associations were observed primarily in prefrontal regions, including frontopolar and inferior frontal areas. **(B) Consumption only.** Correlation maps and significant clusters were derived from studies reporting consumption outcomes only. Significant associations were concentrated in frontopolar and adjacent frontal regions. **(C) Craving and Consumption.** Combined analysis including all eligible craving and consumption outcomes, showing spatial convergence of significant associations across frontal cortical regions. Multiple surface views are provided to facilitate anatomical interpretation.

**Figure 5.**
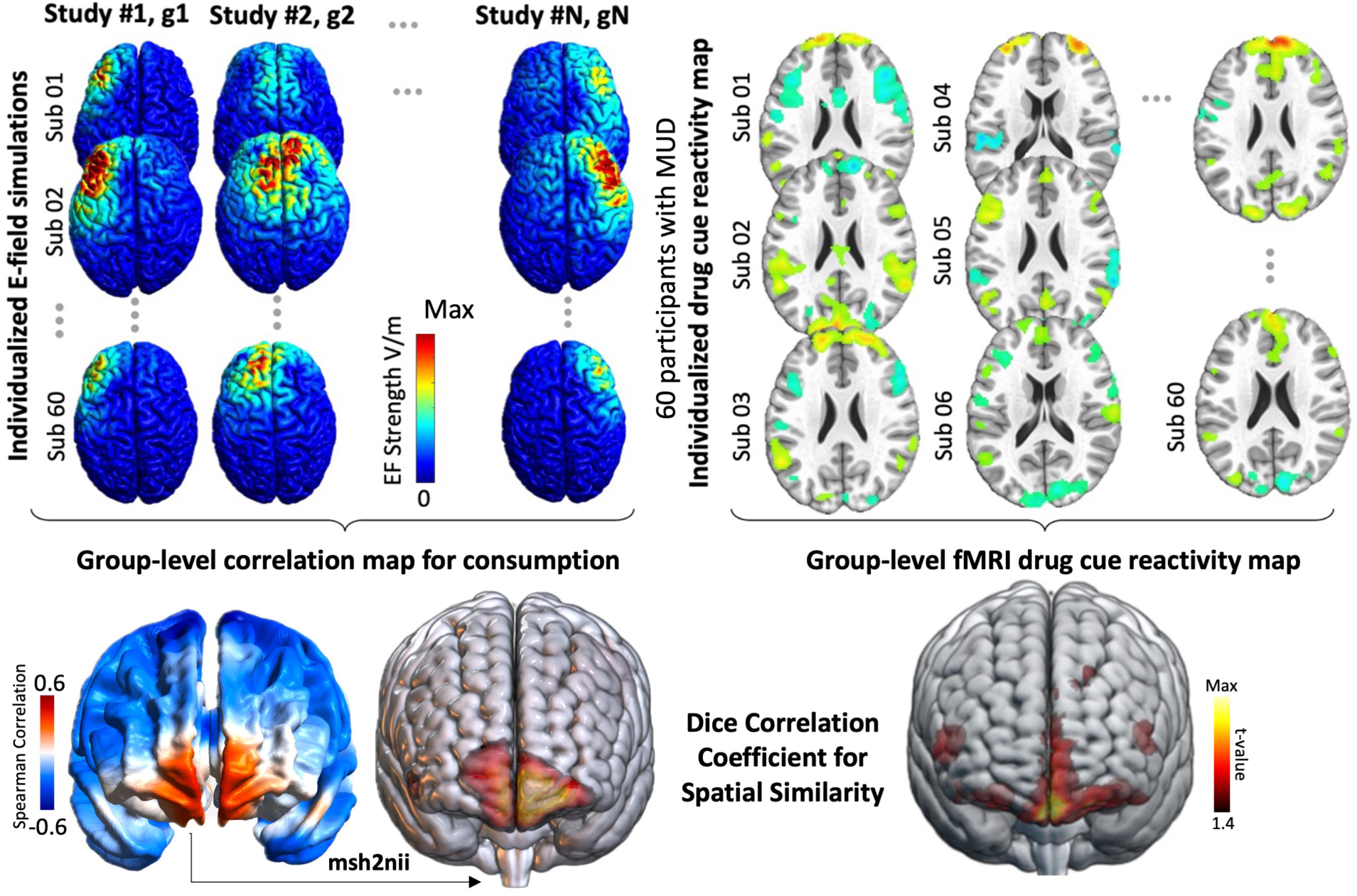
Spatial overlap between the consumption correlation maps and the fMRI drug cue reactivity map. Results are shown for a cohort of 60 individuals with methamphetamine use disorder who completed the structural T1-weighted MRI for individualized computational head modeling and a cue-reactivity fMRI task. The Dice similarity coefficient (DSC) and permutation-based significance testing for the spatial overlap are reported in the figure.

### Primary Results: E-field-Effect Size Associations

#### Consumption-Specific Associations

When analyses were restricted to consumption-related outcomes, node-wise correlation values ranged from r = −0.414 to r = 0.355 (mean ± SD = −0.038 ± 0.078). The 99.9th percentile of positive correlations was r = 0.303. The strongest positive correlation (r = 0.355) was localized to the ventromedial/orbitofrontal region (MNI [−13.6, 67.7, −12.4]), whereas the most negative correlation (r = −0.414) was observed in the medial frontal cortex (MNI [−0.6, −17.7, 4.1]). Following permutation-based multiple-comparison correction, a significant positive cluster was identified over the bilateral frontopolar cortex. No negative clusters survived correction.

#### Craving-Specific Associations

When analyses were restricted to craving-related outcomes, node-wise correlation values ranged from r = −0.195 to r = 0.353 (mean ± SD = 0.081 ± 0.092). The 99.9th percentile of positive correlations was r = 0.300. The strongest positive correlation (r = 0.353) was localized to the left inferior frontal gyrus (IFG; MNI [−50.9, 45.0, −3.2]), whereas the most negative correlation (r = −0.195) was observed in the right dorsolateral prefrontal cortex (MNI [34.3, 19.3, 7.4]). Following permutation-based multiple-comparison correction, two significant clusters were identified: one in the left IFG and another in the left superior frontal gyrus.

### Secondary Results I: Combined Outcome Map

#### Combined Craving and Consumption

As a secondary, descriptive analysis, craving and consumption outcomes were combined to generate a single E-field–effect size association map. Node-wise correlation values ranged from r = −0.176 to r = 0.224 (whole-brain mean ± SD = 0.048 ± 0.061). The strongest positive correlation (r = 0.224) was localized to the left IFG (MNI [−47.7, 48.6, −2.3]), whereas the most negative correlation (r = −0.176) was observed near the medial frontal cortex (MNI [1.1, −1.7, 8.9]). Following permutation-based multiple-comparison correction, two clusters survived statistical significance: a frontopolar/left IFG cluster and a smaller cluster in the left superior frontal gyrus. This combined analysis is presented for descriptive comparison across outcomes and is not interpreted as reflecting a unified biological mechanism.

### Robustness and Sensitivity Analysis

#### Replication of the Results

Additional results obtained without E-field normalization and stratified by coil type (conventional vs. deep) are provided in Supplementary Section S15. Moreover, see Supplementary Section S16 for the replicated results in “Ernie”, a sample healthy subject, and Section S17 for the replication of the results for 60 MUD participants at the group level. We also evaluated the potential influence of the stimulation protocol on the observed E-field–outcome associations (see Supplementary Section S18). We also performed a sensitivity analysis restricted to studies targeting the left DLPFC to evaluate whether the observed E-field–outcome associations were driven by target-location heterogeneity (see Supplementary Section S19).

#### Leave-One-Out Stability Analysis

We evaluated the robustness of the results using leave-one-out (LOO) cross-validation, in which one study was removed at a time, and the analysis was repeated. The resulting maps were highly similar to the full-sample map (mean Pearson r = 0.996; range = 0.928–1.000), indicating that the overall spatial pattern remained stable when individual studies were excluded. Even in the least stable case (r = 0.928), the location of the main peak remained unchanged. Similar levels of stability were observed for both craving and consumption analyses, as well as across different stimulation conditions. In addition, removing individual studies had only minimal effects on the correlation values (median change < 0.002), and results were not related to the effect size of the removed study. Overall, these findings show that the results are robust and not driven by any single study (see Supplementary Section S20 for more details). Additional K-fold resampling analyses (K = 5 and K = 10) demonstrated similarly high spatial reproducibility of the E-field–outcome association maps (mean r = 0.94–0.97; Supplementary Section S21).

#### Sensitivity Analysis for Within-Study Dependence

To assess the influence of multiple effect sizes per study, we conducted a sensitivity analysis using one averaged effect size per study. The resulting spatial correlation maps were highly consistent with the primary analysis (Pearson r = 0.93; Spearman r = 0.93), with identical peak locations across analyses. Although the overlap of the top 1% of elements was moderate (Dice = 0.36), the overall spatial pattern remained highly stable, indicating that the findings are robust to study-level aggregation.

#### Secondary Results II: Spatial Overlap with FDCR

To evaluate spatial correspondence, we computed the Dice similarity coefficient between E-field–effect size association maps and fMRI drug cue–reactivity (FDCR) maps in 60 individuals with MUD (see Supplementary Section S17). The positive consumption-related cluster showed statistically significant spatial correspondence with FDCR activation patterns (Dice coefficient = 0.37). This result reflects spatial consistency between E-field–behavior associations and cue-reactivity–related neural activity and should not be interpreted as validation of causal targets or individualized treatment efficacy.

#### Threshold Robustness

To evaluate the robustness of spatial overlap findings to threshold selection, Dice similarity analyses were repeated across multiple percentile-based thresholds applied to both E-field–effect size association maps and FDCR maps (top 1%, 5%, and 10% of values). Although Dice coefficients decreased with more liberal thresholds (0.37, 0.28, and 0.21 for the top 1%, 5%, and 10%, respectively), the overall pattern of spatial correspondence remained consistent, with statistically significant overlap observed across thresholds. These findings indicate that the observed overlap is not driven by a single threshold choice but reflects consistent spatial alignment between E-field–behavior association maps and cue-reactivity–related neural activity.

## Discussion

Using a multimodal meta-modeling framework, the primary analysis quantified associations between study-level treatment effect sizes and simulated E-field distributions across cortical locations. As contextual support, conventional meta-analysis across 81 randomized TMS trials (107 effect size estimates) showed small-to-moderate improvements in craving and consumption, alongside substantial between-study heterogeneity. Extending beyond this, the primary E-field–effect size analyses showed that higher local E-field strength in prefrontal regions, particularly the bilateral frontopolar cortex for consumption and the left IFG and pre-SMA for craving, was associated with larger symptom reductions across studies. These outcome-specific association maps remained robust in leave-one-out analyses and in sensitivity analyses using one effect size per study, supporting the stability of the observed study-level patterns. Secondary analyses provided complementary but more descriptive evidence: the combined outcome map showed partially overlapping spatial patterns across craving and consumption without implying a unified biological mechanism, and the fMRI overlap analysis showed significant spatial correspondence between the consumption-related E-field map and cue-reactivity–related neural activity in individuals with methamphetamine use disorder. This spatial correspondence remained consistent across threshold levels and should be interpreted as supportive external consistency rather than validation of causal targets or individualized treatment efficacy. Together, these findings provide a quantitative, study-level framework for characterizing how variability in stimulation-induced E-fields relates to variability in observed behavioral outcomes.

While our systematic review showed that the dominant target in previously published papers was left DLPFC (8), our meta-modeling analysis identified three primary cortical regions, frontopolar cortex, IFG, and pre-SMA, where stronger TMS-induced E-fields were consistently associated with greater reductions in craving and/or substance consumption. These regions map onto three core addiction-related neural circuits. The frontopolar area is a central node within fronto-limbic circuitry linking cortical control systems to subcortical reward and salience structures such as the amygdala, ventral striatum, and anterior cingulate cortex (21, 22). In parallel, IFG, a key component of the ventrolateral prefrontal cortex, plays a well-established role in response inhibition, salience attribution, and the regulation of craving (23, 24). Dysfunction within the IFG and its connectivity with striatal and limbic regions have been repeatedly linked to impulsivity, cue-induced craving, and relapse across substance use disorders (25, 26).

Converging anatomical, computational, and clinical evidence further highlights the pre-SMA as a critical link between reward processing and action selection. Located in the medial frontal cortex, the pre-SMA has strong connections with basal ganglia circuits via the hyperdirect pathway, positioning it as a key interface between cortical control systems and striatal pathways involved in compulsive behavior (27). Functionally, it supports action selection and inhibition, and its activity has been associated with impulsivity and addiction severity across disorders (28, 29). Causal studies show that disrupting pre-SMA function reduces sensitivity to reward magnitude and biases decisions toward immediate rewards(30). Consistent with this, neuromodulation studies using theta-burst stimulation (TBS) demonstrate that targeting the pre-SMA can reduce craving and addiction severity, suggesting that this region is functionally dysregulated in addiction rather than simply underactive(31).

These findings suggest that the frontopolar cortex, IFG, and pre-SMA are key nodes within distributed control and reward-related networks implicated in addiction, and that variability in local E-field engagement of these regions is associated with variability in behavioral outcomes across studies. These retrospective associations should not be interpreted as evidence that stimulation of these regions directly causes symptom improvement. Rather, the present findings indicate that greater E-field engagement of these regions tended to co-occur with larger treatment effects across studies. Within this framework, E-field–informed approaches may help refine hypotheses about target selection and dosing by linking stimulation parameters to observed behavioral effects at the study level. Future prospective studies are needed to directly test the causal relevance of these regions by systematically manipulating stimulation targets, E-field magnitude, and stimulation parameters while measuring subsequent changes in craving and substance use outcomes. Such studies could determine whether intentionally increasing E-field engagement in these candidate regions leads to improved clinical outcomes and thereby validate the hypotheses generated by the present meta-modeling framework.

Inspired by prior work using symptom-specific meta-modeling approaches in schizophrenia (17) and related neuropsychiatric conditions (32), our outcome-specific correlation maps suggest both shared and dissociable patterns of association across craving and consumption. Although both outcomes were linked to prefrontal control regions, craving-related effects were more strongly associated with lateral prefrontal areas, including IFG, consistent with their established roles in salience processing, inhibitory control, and cue-induced motivation. In contrast, consumption-related effects showed stronger associations with the frontopolar cortex, a region implicated in long-term goal maintenance and sustained behavioral regulation. These patterns suggest that prior TMS studies targeting craving versus consumption may have engaged partially distinct cortical systems and E-field distributions.

Extending these findings, we observed statistically significant spatial correspondence between the consumption-related E-field–effect size association map and the FDCR activation map in a clinical sample of individuals with MUD. This correspondence indicates that cortical regions where higher TMS-induced E-fields were associated with larger reductions in substance consumption overlap with neural circuits engaged during drug cue processing. This observation is consistent with prior work reporting that spatial overlap between E-field distributions and cue-reactivity maps is associated with variability in treatment outcomes in individuals with SUD receiving TMS over the frontopolar cortex (33). These findings suggest that medial prefrontal regions, particularly the frontopolar cortex, may represent candidate loci where stimulation-induced E-fields align with neural systems implicated in cue-reactivity and behavioral control, consistent with prior work implicating the frontopolar cortex in prospective decision-making and long-term goal maintenance (34, 35).

## Limitation and Future Directions

Despite the robustness of the findings, several limitations should be considered. (1) Heterogeneity in outcome assessment, including differences in measurement instruments and timing for craving and consumption, represents an important source of variability across studies, despite efforts to harmonize outcomes. (2) The meta-modeling framework identifies study-level associations between E-field distributions and treatment outcomes and therefore should not be interpreted as providing causal evidence or definitive stimulation targets. (3) The spatial overlap analysis was conducted in a single MUD cohort and should be considered supportive rather than a validation of the broader framework. (4) Although stimulation intensity was incorporated into E-field modeling, other dimensions of TMS dose (e.g., pulse number, treatment duration, and session scheduling) were not explicitly modeled. (5) The analysis was restricted to study-level target definitions and effect sizes, limiting the ability to evaluate individualized targeting strategies and participant-level dose–response relationships. Detailed discussion of these limitations and future directions is provided in Supplementary Section S22.

## Conclusion

A major strength of this study is the explicit modeling of TMS-induced E-fields to relate stimulation parameters to behavioral effect sizes at the study level. This framework provides a quantitative approach for examining how variability in induced E-fields corresponds to variability in observed outcomes across studies. The results were robust across multiple analyses, including replication in an independent clinical sample using anatomically individualized head models and consistent spatial correspondence with cue-reactivity–related neural activity. In conclusion, the present findings indicate that local E-field strength in the frontopolar cortex, IFG, and pre-SMA is associated with variability in craving and substance consumption outcomes across studies of substance use disorders. These results are hypothesis-generating and provide a basis for future studies to further investigate the relationship between stimulation-induced fields and behavioral outcomes in clinical trials.

## Data Availability

All data produced in the present study are available upon reasonable request to the authors

## Authors’ Contribution

G.S. contributed to the conception and design of the study, systematic review update, data extraction, electric-field simulations, statistical analyses, interpretation of findings, and drafting of the manuscript. H.E. contributed to the interpretation of clinical and translational relevance, revision of the manuscript, and provided fMRI data. A.O. contributed to study conception and design, methodological development of electric-field modeling and meta-modeling framework, supervision of analyses, interpretation of results, and critical manuscript revision. All authors approved the final version of the manuscript and agree to be accountable for all aspects of the work.

## Acknowledgments

G.S. was supported by a fellowship from the University of Minnesota’s MnDRIVE (Minnesota’s Discovery, Research and Innovation Economy) initiative. No generative artificial intelligence tools were used in the design of the study, data analysis, interpretation of results, or preparation of the scientific content of the manuscript.

## Code and Data availability

Detailed data extraction can be accessed at https://osf.io/sv8ky/overview. Simulation results for a representative standard subject are available upon request. All analysis code will be released in a GitHub repository following manuscript acceptance.

## References

1. Mateu-Mollá J, Pérez-Gálvez B, Villanueva-Blasco VJ. Pharmacological treatment for substance use disorder: A systematic review. Addictive behaviors. 2025:108242.

2. Bormann NL, Oesterle TS, Arndt S, Karpyak VM, Croarkin PE. Systematic review and meta-analysis: Combining transcranial magnetic stimulation or direct current stimulation with pharmacotherapy for treatment of substance use disorders. The American Journal on Addictions. 2024;33(3):269–82.

3. Siebner HR, Funke K, Aberra AS, Antal A, Bestmann S, Chen R, et al. Transcranial magnetic stimulation of the brain: What is stimulated?–A consensus and critical position paper. Clinical neurophysiology. 2022;140:59–97.

4. Kearney-Ramos TE, Dowdle LT, Lench DH, Mithoefer OJ, Devries WH, George MS, et al. Transdiagnostic effects of ventromedial prefrontal cortex transcranial magnetic stimulation on cue reactivity. Biological Psychiatry: Cognitive Neuroscience and Neuroimaging. 2018;3(7):599–609.

5. Hanlon CA, Dowdle LT, Gibson NB, Li X, Hamilton S, Canterberry M, et al. Cortical substrates of cue-reactivity in multiple substance dependent populations: transdiagnostic relevance of the medial prefrontal cortex. Translational psychiatry. 2018;8(1):186.

6. Deng Z-D, Lisanby SH, Peterchev AV. Electric field depth–focality tradeoff in transcranial magnetic stimulation: simulation comparison of 50 coil designs. Brain stimulation. 2013;6(1):1–13.

7. Opitz A, Windhoff M, Heidemann RM, Turner R, Thielscher A. How the brain tissue shapes the electric field induced by transcranial magnetic stimulation. Neuroimage. 2011;58(3):849–59.

8. Soleimani G, Souki A, Honari S, Baker TE, Brunoni AR, Ebrahimi M, et al. Effectiveness of Noninvasive Brain Stimulation Protocols on Drug Craving and Consumption/Relapse in Substance Use Disorders: A Systematic Review and Meta-analysis of 208 Clinical Trials and 36 Protocols. medRxiv. 2025:2025.09. 21.25335559.

9. Mehta DD, Praecht A, Ward HB, Sanches M, Sorkhou M, Tang VM, et al. A systematic review and meta-analysis of neuromodulation therapies for substance use disorders. Neuropsychopharmacology. 2023:1–32.

10. Nieminen JO, Koponen LM, Ilmoniemi RJ. Experimental characterization of the electric field distribution induced by TMS devices. Brain stimulation. 2015;8(3):582–9.

11. Drakaki M, Mathiesen C, Siebner HR, Madsen K, Thielscher A. Database of 25 validated coil models for electric field simulations for TMS. Brain stimulation. 2022;15(3):697–706.

12. Laakso I, Hirata A, Ugawa Y. Effects of coil orientation on the electric field induced by TMS over the hand motor area. Physics in Medicine & Biology. 2013;59(1):203.

13. Caulfield KA, Brown JC. The problem and potential of TMS’infinite parameter space: a targeted review and road map forward. Frontiers in Psychiatry. 2022;13:867091.

14. Wischnewski M, Mantell KE, Opitz A. Identifying regions in prefrontal cortex related to working memory improvement: A novel meta-analytic method using electric field modeling. Neuroscience & Biobehavioral Reviews. 2021;130:147–61.

15. Wischnewski M, Berger TA, Opitz A, Alekseichuk I. Causal functional maps of brain rhythms in working memory. Proceedings of the National Academy of Sciences. 2024;121(14):e2318528121.

16. Wischnewski M, Berger TA, Opitz A. Meta-modeling the effects of anodal left prefrontal transcranial direct current stimulation on working memory performance. Imaging Neuroscience. 2024;2:1–14.

17. Sinanaj L, Pallis K, Dehkordi AF, Huguelet P, Kaiser S, Bègue I. Mapping symptom-general and symptom-specific targets for transcranial magnetic stimulation in schizophrenia: an electric-field modeling meta-analysis. Molecular Psychiatry. 2025:1–11.

18. Page MJ, McKenzie JE, Bossuyt PM, Boutron I, Hoffmann TC, Mulrow CD, et al. The PRISMA 2020 statement: an updated guideline for reporting systematic reviews. Bmj. 2021;372.

19. Thielscher A, Antunes A, Saturnino GB, editors. Field modeling for transcranial magnetic stimulation: a useful tool to understand the physiological effects of TMS? 2015 37th annual international conference of the IEEE engineering in medicine and biology society (EMBC); 2015: IEEE.

20. Baetens K, Van Hoornweder S, Berger TA, Wischnewski M. ACES: Automated Correlation of Electric field strength and Stimulation effects for non-invasive brain stimulation. Brain Stimulation: Basic, Translational, and Clinical Research in Neuromodulation. 2024;17(2):473–5.

21. Jasinska AJ, Chen BT, Bonci A, Stein EA. Dorsal medial prefrontal cortex (MPFC) circuitry in rodent models of cocaine use: implications for drug addiction therapies. Addiction biology. 2015;20(2):215–26.

22. Goldstein RZ, Volkow ND. Dysfunction of the prefrontal cortex in addiction: neuroimaging findings and clinical implications. Nature reviews neuroscience. 2011;12(11):652–69.

23. Qiu Z, Wang J. Altered neural activities during response inhibition in adults with addiction: a voxel-wise meta-analysis. Psychological medicine. 2021;51(3):387–99.

24. Swick D, Ashley V, Turken. Left inferior frontal gyrus is critical for response inhibition. BMC neuroscience. 2008;9(1):102.

25. Dakhili A, Sangchooli A, Jafakesh S, Zare-Bidoky M, Soleimani G, Batouli SAH, et al. Cue-induced craving and negative emotion disrupt response inhibition in methamphetamine use disorder: Behavioral and fMRI results from a mixed Go/No-Go task. Drug and Alcohol Dependence. 2022;233:109353.

26. Viswanath H, Velasquez KM, Savjani R, Molfese DL, Curtis K, Molfese PJ, et al. Interhemispheric insular and inferior frontal connectivity are associated with substance abuse in a psychiatric population. Neuropharmacology. 2015;92:63–8.

27. Eickhoff SB, Bzdok D, Laird AR, Roski C, Caspers S, Zilles K, et al. Co-activation patterns distinguish cortical modules, their connectivity and functional differentiation. Neuroimage. 2011;57(3):938–49.

28. Lohse A, Løkkegaard A, Siebner HR, Meder D. Linking impulsivity to activity levels in pre-supplementary motor area during sequential gambling. Journal of Neuroscience. 2023;43(8):1414–21.

29. Huang D, Ma Y-Y. Increased excitability of layer 2 cortical pyramidal neurons in the supplementary motor cortex underlies high cocaine-seeking behaviors. Biological psychiatry. 2023;94(11):875–87.

30. Vural G, Katruss N, Soutschek A. Pre-supplementary motor area strengthens reward sensitivity in intertemporal choice. NeuroImage. 2024;299:120838.

31. Pallanti S, Di Ponzio M, Levola J, Lioumis P, Paunio T, Kičić D, et al. Treatment of behavioral addictions and substance use disorders: A focus on the effects of theta-burst stimulation over the pre-SMA. International Journal of Mental Health and Addiction. 2025;23(4):2808–21.

32. Siddiqi SH, Taylor SF, Cooke D, Pascual-Leone A, George MS, Fox MD. Distinct symptom-specific treatment targets for circuit-based neuromodulation. American Journal of Psychiatry. 2020;177(5):435–46.

33. McCalley DM, Hanlon CA. The importance of overlap: a retrospective analysis of electrical field maps, alcohol cue-reactivity patterns, and treatment outcomes for alcohol use disorder. Brain Stimulation: Basic, Translational, and Clinical Research in Neuromodulation. 2023;16(3):724–6.

34. Soleimani G, Joutsa J, Moussawi K, Siddiqi SH, Kuplicki R, Bikson M, et al. Converging Evidence for Frontopolar Cortex as a Target for Neuromodulation in Addiction Treatment. American Journal of Psychiatry. 2023:appi. ajp. 20221022.

35. Joutsa J, Moussawi K, Siddiqi SH, Abdolahi A, Drew W, Cohen AL, et al. Brain lesions disrupting addiction map to a common human brain circuit. Nature medicine. 2022;28(6):1249–55.

