## Supplementary Materials for "Associations Between TMS-Induced Electric Fields and Craving and Consumption Outcomes in Substance Use Disorders: A Multimodal Dose-Response Meta-Analysis"

**Title:**

**Running Title:**

Electric Field-Behavior Associations in Addiction

**Authors and Affiliations**

Ghazaleh Soleimani (PhD)[1,#], Martin P Paulus (MD)[2], Hamed Ekhtiari (MD-PhD)[3], Alexander Opitz (PhD)[1,#]

[1] Department of Psychiatry and Behavioral Sciences, University of Minnesota, MN, USA
[2] Laureate Institute for Brain Research (LIBR), OK, USA
[3] Department of Psychiatry, University of Texas Southwestern (UTSW), TX, USA.

### **Corresponding Authors**

Ghazaleh Soleimani
University of Minnesota

Alexander Opitz
University of Minnesota

**Key words:**
Substance use disorder, Meta modeling, Electric field, Transcranial Magnetic stimulation (TMS), Drug cue-reactivity

**S1. Example of Heterogeneity in TMS Targeting Approaches**

For example, studies aiming to modulate the executive control network commonly position the TMS coil over the F3 location of the international EEG system (e.g., (1, 2, 3)) to target the left dorsolateral prefrontal cortex (DLPFC). In contrast, other investigations have used alternative scalp locations (e.g., F5(4)) or non–EEG-based targeting methods, including fixed scalp-distance rules (e.g., 5 cm(5, 6, 7) or 6 cm(8, 9, 10) rules), the Beam F3 method (e.g., (11, 12)), or individualized functional (e.g., (13, 14)) or structural MRI-guided (e.g., neuronavigation(15, 16)) approaches. Because targeting strategies directly shape the spatial distribution of the induced E-field, heterogeneity in target selection represents a particularly important source of inter-experimental variability(17). However, conventional meta-analytic approaches typically average across heterogeneous stimulation features and therefore do not explicitly model how variation in stimulation parameters shapes the spatial distribution of the induced E-field and contributes to variability in observed treatment effect sizes.

**S2. Search Strategy for Data Extraction**

A comprehensive literature search was performed using the PubMed database to identify all published studies up to the end of 2025 that applied TMS for substance use disorders (SUDs). Key terms related to transcranial magnetic stimulation (TMS) included “transcranial magnetic stimulation,” TMS, “repetitive transcranial magnetic stimulation,” rTMS, “theta burst stimulation,” TBS, “intermittent theta burst stimulation,” iTBS, “continuous theta burst stimulation,” cTBS, “deep transcranial magnetic stimulation,” dTMS, “high-frequency transcranial magnetic stimulation,” “low-frequency transcranial magnetic stimulation,” and “magnetic stimulation.” Key terms related to addiction included addiction, “substance use,” “substance abuse,” “substance-related disorder,” “drug abuse,” “behavioral addiction,” dependency, craving, alcohol, nicotine, tobacco, smoking, cigarette, cannabis, opioid, marijuana, crack, cocaine, morphine, heroin, and methamphetamine. In each study, different factors that can affect stimulation outcomes and E-field distribution patterns were extracted (Supplementary Table S1).

This search yielded 81 TMS studies eligible for inclusion in the systematic review. Studies were included if they reported at least one outcome measure related to craving or consumption and provided sufficient data to compute effect sizes for meta-analysis. Across the 81 included studies, a total of 107 outcome measures were reported, comprising 75 craving-related and 32 consumption-related outcomes. Effect sizes were calculated using Comprehensive Meta-Analysis software (version 4; Englewood, NJ, USA), focusing on quantifying TMS-induced changes in craving or substance use. For each study, the following data were extracted: outcome variables required to compute Hedges’ g values (with positive values indicating reductions in craving or consumption and thus clinical improvement), sample sizes in active and sham groups, type of SUD, stimulation target location, coil type, stimulation intensity, stimulation duration, and number of sessions. A detailed overview of all extracted variables is provided in Table S1.

The present meta-modeling analysis builds upon a previously conducted comprehensive systematic review and meta-analysis of non-invasive brain stimulation in substance use disorders (Soleimani et al., 2025, pre-registered in OSF https://osf.io/sv8ky/overview; however, no protocol was prospectively registered for the present secondary analysis), which has been updated by the end of 2025, followed by PRISMA guidelines for study identification, screening, eligibility assessment, and data extraction. The current study extends the original review by incorporating electric-field simulation and correlation-based meta-modeling.

For the current study, the existing curated database was updated through an additional PubMed search extending to December 2025 using the same search strategy and eligibility criteria. Screening and primary data extraction procedures for studies included in the original review were performed according to PRISMA standards and are described in detail in the prior publication. For the present meta-modeling analysis, additional study-level variables relevant to electric-field simulation (e.g., stimulation target coordinates, coil type, orientation, and stimulation intensity) were extracted using a standardized form. When required parameters were not explicitly reported, standardized assumptions based on validated simulation templates were applied.

**S3. Inclusion/Exclusion Criteria**

The initial literature search identified 1,120 records. Following title and abstract screening, 610 TMS-related articles were excluded. Exclusion criteria at this stage were book chapters, commentaries, author corrections, editorials, and studies focusing on conditions other than SUDs. The remaining 510 articles were assessed in full for eligibility. Full-text review resulted in the inclusion of 81 TMS studies, comprising 107 distinct clinical outcome measures. Articles were excluded at this stage if they were review papers, case reports, or case series, study protocols, or non-human studies; if they enrolled only healthy participants; if they focused solely on E-field modeling; if they lacked sufficient methodological detail on stimulation montages; or if they were published in languages other than English.

**S4. Meta-Analysis Approach, Effect size extraction, and Risk of Bias Assessment**

Studies identified in the systematic review were considered for inclusion in the meta-analysis based on predefined eligibility criteria. To be included, studies were required to: (1) report relevant outcomes, including changes in substance craving, consumption, abstinence, or relapse; and (2) provide sufficient data to compute effect sizes, including either (i) means and standard deviations (or standard errors) for both active and sham stimulation groups before and after intervention, (ii) the mean difference and associated standard deviation (or standard error) between active and sham groups, or (iii) reported P or F values for comparisons between active and sham groups. Studies were excluded from the meta-analysis if they: (1) did not specifically examine the effects of neuromodulation on SUDs, particularly with respect to craving, consumption, relapse, or abstinence; or (2) lacked essential statistical information required for effect size calculation. In cases of missing data, the first or last authors were contacted to request additional information; studies were excluded if the required data could not be obtained or if no response was received.

Meta-analysis was conducted using the “metafor” package in R. Standardized effect sizes were computed by contrasting changes in behavioral measures between active stimulation and sham control arms. All outcomes were summarized using Hedges’ g with corresponding 95% confidence intervals. To accommodate heterogeneity across study designs and populations, random-effects modeling was employed. For studies reporting both craving and consumption outcomes, a separate Hedges' g was computed for each outcome to represent the effect size associated with the applied TMS. Positive Hedges' g values denote beneficial effects of TMS, whereas values at or below zero indicate no measurable improvement. For studies reporting multiple Hedges' g values for the same TMS protocol (e.g., from distinct craving questionnaires or timepoints), a single summary effect size per outcome was computed to maintain statistical independence. Effect sizes were combined using inverse-variance weighting when variance data were available; otherwise, unweighted arithmetic means were calculated.

Assessment of study reliability and risk of bias was conducted as part of the main INTAM meta-analysis pipeline (18). Briefly, risk of bias was evaluated using the second version of the Cochrane Risk of Bias tool for randomized trials (RoB 2), which assesses five domains: the randomization process, deviations from intended interventions, missing outcome data, measurement of outcomes, and selection of reported results. Each domain was rated as "low risk," "some concerns," or "high risk." An initial calibration phase was performed in which five randomly selected studies were jointly reviewed to resolve ambiguities and ensure consistency. Subsequently, two independent reviewers assessed the risk of bias for each included study.

**S5. Electric Field Modeling for the SUD Population**

To generate anatomically realistic E-field maps for the SUD population, reported MNI stimulation coordinates from each study were transformed into individual subject space using the "*mni2subject_coords*" function implemented in SimNIBS. E-field simulations were then performed for each of the 60 head models, applying study-specific coil parameters across all experimental conditions. Tissue-specific conductivity parameters were incorporated into each subject model to enable personalized E-field estimation (19). Conductivities for scalp, skull, and gray matter were sampled from a β(3,3) distribution to represent biological uncertainty (20, 21), while values for cerebrospinal fluid, white matter, and ocular tissues were fixed given their negligible variability effects. Conductivity ranges included $\sigma_{skull}$ = 0.002–0.03 S/m, $\sigma_{skin}$ = 0.2–0.6 S/m, $\sigma_{gray}$ = 0.1–0.6 S/m, $\sigma_{CSF}$ = 1.66 S/m, $\sigma_{eye}$ = 0.5 S/m, and $\sigma_{white\ matter}$ = 0.14 S/m.

**S6. Imaging Parameters for the SUD Population**

Structural and functional MRIs were obtained on two identical GE MRI 750 3T scanners. Structural MRI parameters: TR/TE = 5/2.012 ms, FOV/slice = 24 x 192/0.9 mm, 256x256 matrix producing 0.938 x 0.9 mm voxels and 186 axial slices for T1-weighted images and TR/TE=8108/137.728ms, FOV/slice=240/2mm, 512x512 matrix producing 0.469x0.469x2mm voxels and 80 coronal slices for T2-weighted images. T1- and T2-weighted MR images were used to generate computational head models for each individual. Task-state fMRI parameters: TR/TE = 2000/27 ms, FOV/slice = 240/2.9 mm, 128x128 matrix producing 1.857x1.857x2.9 mm voxels, 39 axial slices, and 196 repetitions.

**S7. Demographic Data for the SUD Population**

Sixty participants with methamphetamine use disorder (MUD) were enrolled in the study. All participants were male, with a mean age ± standard deviation of 35.86 ± 8.47 years (range: 20–

55 years). Participants were recruited during early abstinence from the 12&12 Residential Drug Addiction Treatment Center in Tulsa, Oklahoma. All human research procedures were conducted in accordance with the Declaration of Helsinki and applicable ethical guidelines and regulations. Written informed consent was obtained from all participants before study procedures, and the study protocol was approved by the Western Institutional Review Board (WIRB Protocol #20171742).

**Table S2. Demographics and substance use profile.** All 60 participants are diagnosed with MUD

| **Demographic data** | Mean (SD) |
|---|---|
| Age (years) | 36.8 (8.7) |
| Education (years) | 13.6 (2.6) |
| BMI | 28.0 (5.1) |
| Age of Meth use onset (years old) | 20.7 (7.1) |
| Duration of Meth use at least once a week (years) | 13.2 (19.8) |
| Cost of Meth (dollar per month) | 1039.5 (1267.1) |
| Dose of Meth (gram per day) | 1.61 (1.51) |
| Life time history of meth injection [b] (n (%)) | 46 (77%) |
| Days of drug use in the last month (before starting abstinence) | |
| Meth | 22.4 (10.0) |
| Alcohol | 9.4 (11.7) |
| Heroin | 3.9 (9.1) |
| Methadone | 0.7 (3.9) |
| Other opioids | 2.6 (6.1) |
| Barbiturate | 0.1 (0.6) |
| Sedative | 2.2 (6.2) |
| Cocaine | 0.2 (0.9) |
| Cannabis | 7.2 (9.8) |
| Hallucinogens | 0.4 (1.9) |
| Inhalants | 0.1 (0.4) |
| Duration of current abstinence (days) | 61.5 (34.8) |
| Craving Changes on VAS (0-100) from before to after Scanning | 20.8 (25.8) |

#### S8. fMRI Task Details and Processing

Task-based fMRI data were acquired during a pictorial methamphetamine cue-exposure paradigm using a pseudo-randomized block design. The task included two stimulus categories—methamphetamine-related and neutral images—drawn from two distinct but equivalent image sets that were previously validated (22). Each block comprised six images from the same category (meth or neutral), with each image presented for 5 s and separated by a 0.2 s interstimulus interval. Between blocks, a fixation cross was displayed for a jittered duration of 8–12 s. The task consisted of four methamphetamine-related blocks and four neutral blocks (eight blocks total), resulting in six images per category per block. The total task duration was approximately 6 min. The fMRI task code and stimulus materials are publicly available at https://github.com/rkuplicki/LIBR_MOCD.

Functional activity analysis was performed in AFNI. The first three pre-steady state images were removed, and preprocessing steps were despiking, slice timing correction, realignment, transformation to MNI space, and 4 mm of Gaussian FWHM smoothing. Three polynomial terms and the six motion parameters were regressed out. TRs with excessive motion (defined as Euclidean norm of derivative of the six motion parameters being greater than 0.3) were censored during regression. Single-subject general linear models (GLMs) were constructed using AFNI's "3dDeconvolve". Methamphetamine-related and neutral cue blocks were modeled as separate regressors of interest and convolved with a canonical hemodynamic response function. Beta coefficients were estimated for each condition, and subject-level contrast maps were generated by computing the methamphetamine > neutral cue contrast to index drug cue reactivity. Resulting contrast images were carried forward to group-level analyses. Group-level statistical analyses were performed using AFNI's mixed-effects modeling framework, accounting for between-subject variability. Statistical significance was assessed using voxelwise thresholds combined with cluster-level correction for multiple comparisons, as implemented in AFNI "3dClustSim", to control the family-wise error rate. For visualization and comparison to correlation map purposes, the final map was thresholded to display beta coefficients within the range of −3 to 3.

**S9. Statistical Inference and Robustness Analyses Methods**

Permutation-Based Significance Testing: Statistical significance of the E-field–effect size associations was evaluated using a nonparametric permutation framework. Specifically, study-level effect size labels (Hedges' g) were randomly shuffled across studies while preserving the corresponding E-field distributions, thereby generating null distributions under the hypothesis of no association between local E-field strength and behavioral effect sizes. This procedure was repeated 5,000 times to obtain stable empirical null distributions(23).

Multiple-Comparison Correction: To account for multiple comparisons across cortical nodes, a maximum-statistic approach was used. For each permutation, the maximum absolute correlation value across all nodes was retained to construct a family-wise error (FWE)–corrected null distribution. Observed node-wise correlation values were then compared against this distribution to determine statistical significance, controlling for multiple comparisons across the cortical surface.

Leave-One-Out Stability Analysis: To assess the robustness of the findings, leave-one-out (LOO) cross-validation was performed. At each iteration, data from a single study were excluded, and the E-field–effect size association analysis was recomputed using the remaining dataset. This process was repeated across all studies, yielding a set of LOO-derived correlation maps. Map stability was quantified by calculating spatial similarity between each LOO map and the full-sample map. High spatial correspondence and consistent localization of peak regions were interpreted as evidence of robustness to the exclusion of individual studies. For further robustness analyses, K-fold cross-validation (with K=5 and K=10) was conducted as supplementary analyses (see Supplementary Section S15).

Sensitivity Analysis for Within-Study Dependence: To evaluate the potential impact of dependence among multiple effect sizes derived from the same study, sensitivity analyses were conducted using a one-effect-per-study approach. In these analyses, a single effect size was retained for each study, and the full association mapping procedure was repeated. Consistency of results across the primary and sensitivity analyses was used to assess the robustness of findings to within-study dependence.

Threshold-Dependent Spatial Overlap Analysis: Because Dice similarity depends on thresholding of continuous maps, spatial overlap analyses were conducted across multiple threshold levels. Both E-field–effect size maps and FDCR maps were thresholded using percentile-based criteria (top 1%, 5%, and 10% of values), and Dice coefficients were computed for each threshold. Consistency of overlap across thresholds was used to assess the robustness of spatial correspondence and to ensure that results were not driven by a single threshold selection. Statistical significance of Dice values was evaluated using permutation testing, in which one map was randomly rotated or relabeled to generate a null distribution of overlap values.

#### S10. Summarizing data extraction

To provide a comprehensive overview of study characteristics extracted from the included trials, Figure S8 presents the distribution of studies across substance type, stimulation protocol, outcome domain, and assessment instrument. This visualization complements the descriptive summary reported in the main manuscript and offers a detailed representation of methodological heterogeneity across trials.

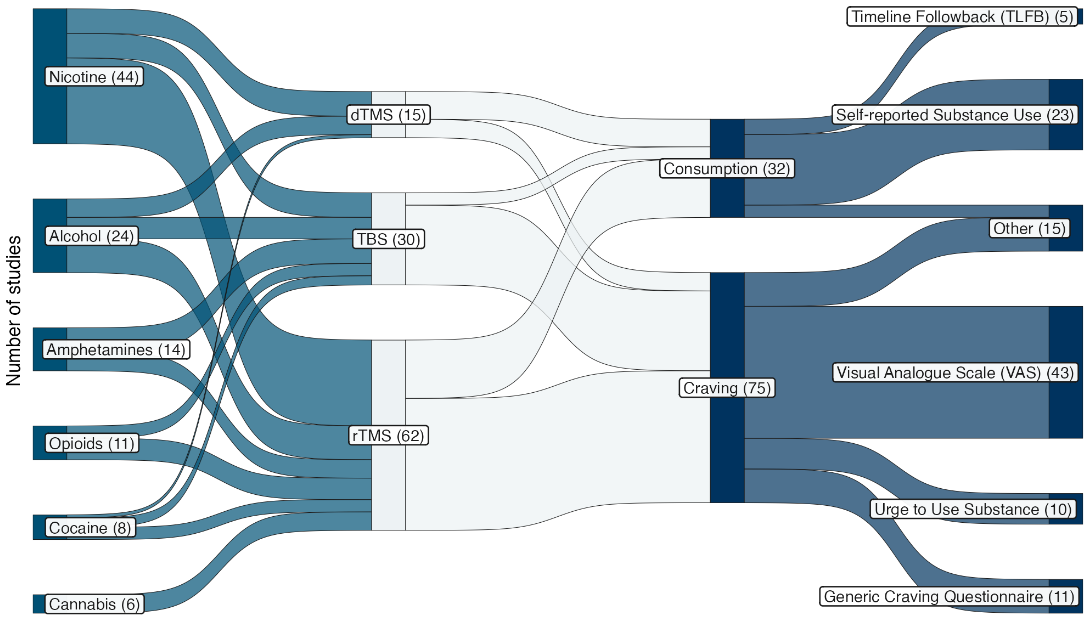


**Figure S1. Distribution of outcome assessments across TMS trials for substance use disorders.** This Sankey diagram illustrates the flow of included studies across four categorical dimensions: substance use disorder (SUD) type, stimulation protocol, primary outcome domain, and assessment instrument. The leftmost column shows the number of studies targeting different SUD populations (nicotine, alcohol, amphetamines, opioids, cocaine, and cannabis). These studies are then grouped by stimulation protocol (rTMS, theta-burst stimulation [TBS], and deep TMS [dTMS]), followed by outcome domain (craving or consumption), and finally by the specific measurement instruments used to quantify outcomes (e.g., visual analogue scale [VAS], self-reported substance use, Timeline Followback [TLFB], urge-to-use scales, and generic craving questionnaires). Box widths within each column represent the relative number of studies in each category, and ribbon widths indicate the proportion of studies shared between categories across adjacent columns, thereby highlighting how stimulation modalities and clinical targets map onto downstream outcome measures and instruments used in the literature.

#### S11. Standard Meta-analysis results

**Substance-specific effects:** When stratified by substance, the largest pooled effects were observed for opioid use disorders (k = 11; g = 1.08, 95% CI [0.42, 1.74]) and amphetamine use disorders (k = 14; g = 0.95, 95% CI [0.59, 1.30]), followed by nicotine (k = 44; g = 0.55, 95% CI [0.24, 0.87]) and cocaine (k = 8; g = 0.54, 95% CI [0.14, 0.94]). Effects for alcohol were smaller and did not reach statistical significance (k = 24; g = 0.24, 95% CI [−0.01, 0.50]), while cannabis studies showed no reliable effect (k = 6; g = 0.16, 95% CI [−0.11, 0.43]). Heterogeneity was high for most substances ($I^2$ = 62–89%), indicating considerable variability across studies.

**Craving versus consumption outcomes:** When outcomes were categorized by type, craving measures demonstrated significantly larger effects than consumption measures. Across all craving outcomes (k = 75), stimulation yielded a moderate-to-large pooled effect (g = 0.64, 95% CI [0.42, 0.86]), with high heterogeneity ($I^2$ = 88%). In contrast, consumption outcomes (k = 32) showed a smaller but significant pooled effect (g = 0.39, 95% CI [0.17, 0.60]), with moderate heterogeneity ($I^2$ = 63%).

**Stimulation parameters:** High-frequency stimulation was associated with larger effects than low-frequency stimulation for both craving (HF: k = 56; g = 0.78, 95% CI [0.51, 1.04]) and consumption (HF: k = 30; g = 0.34, 95% CI [0.13, 0.56]), whereas low-frequency stimulation showed smaller and more variable effects across outcome categories.

#### S12. Electric Field Distribution Details

TMS-induced E-field varied substantially across studies as a function of stimulation parameters, including target location, coil type, orientation, and intensity. Target localization approaches included rule-based methods (e.g., 5-cm rule, Beam F3), EEG 10–20 system coordinates (e.g., F3, F4, Fp1, Fz, AFz), as well as neuronavigation and fMRI-guided targeting. For the latter category, only studies using group-level neuroimaging-derived targets (i.e., a single target coordinate applied consistently across participants) were included; studies employing individualized neuroimaging-guided targeting, where stimulation targets varied across participants based on subject-specific imaging data, were not considered. This approach ensured that study-level stimulation targets could be modeled with reasonable fidelity across studies.

Stimulation intensity was typically defined relative to motor threshold (median = 100%, range = 80–120%). Conventional figure-of-eight coils were most commonly used (67.3%, n = 72), whereas deep TMS protocols represented a smaller subset (13.1%, n = 14); coil type was not reported in 18.7% of studies (n = 20). For studies in which coil type was not explicitly reported, coil models were assigned based on the stimulation device when available. Specifically, studies using Magstim devices were modeled using the Magstim_70mm_Fig8 coil, whereas studies using MagVenture devices were modeled using the MagVenture_Cool-B65 coil. If neither the coil type nor the stimulation device was reported, the Magstim_70mm_Fig8 coil was used as the default model because it was the most commonly used coil across the included studies.

#### S13. Spatial Variability of Electric-Field Distributions Across Common TMS Targets

To further characterize the spatial distribution of stimulation delivered across studies, we generated average E-field maps for the four most frequently targeted cortical regions identified in the systematic review: left dorsolateral prefrontal cortex (Left DLPFC; n = 56), right dorsolateral prefrontal cortex (Right DLPFC; n = 15), medial prefrontal cortex (mPFC; n = 16), and all other locations (including left IFG, right IFG, Fz, Cz, and left parietal) (Figure S8). For each study, E-field maps were scaled according to the reported stimulation intensity (dI/dt) and projected onto a common cortical template. Mean and standard deviation maps were then calculated across studies within each anatomical target category.

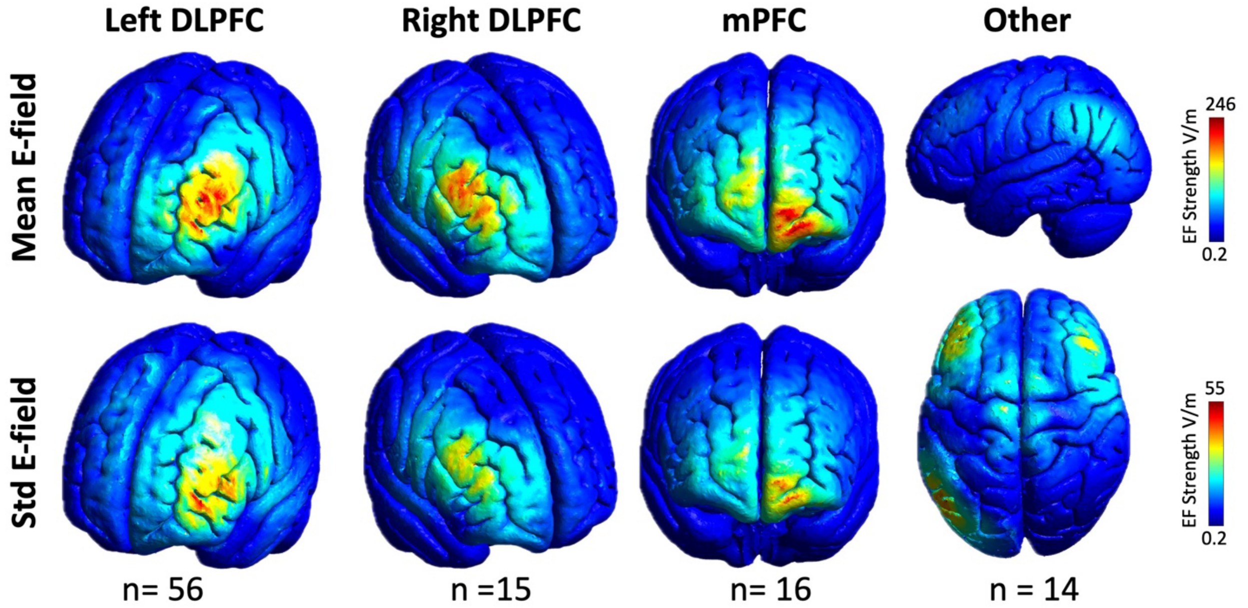


**Figure S2. Mean and standard deviation of induced electric-field (E-field) distributions across the four most commonly targeted cortical regions in the included TMS studies.** The top row shows the mean E-field strength and the bottom row shows the standard deviation of E-field strength across studies targeting the left dorsolateral prefrontal cortex (Left DLPFC), right dorsolateral prefrontal cortex (Right DLPFC), medial prefrontal cortex (mPFC), and all other remaining targets (left and right IFG, Fz, Cz, and left parietal). E-field maps were generated by averaging subject-specific E-field simulations after scaling by the reported stimulation intensity (dI/dt) and projecting them onto a common cortical template. Warmer colors indicate higher E-field strength, whereas cooler colors indicate lower E-field strength.

#### S14. Averaged Electric Field based on Conventional vs Deep Coils

Averaged E-field values divided by coil types (conventional vs deep) are provided in Figure S1.

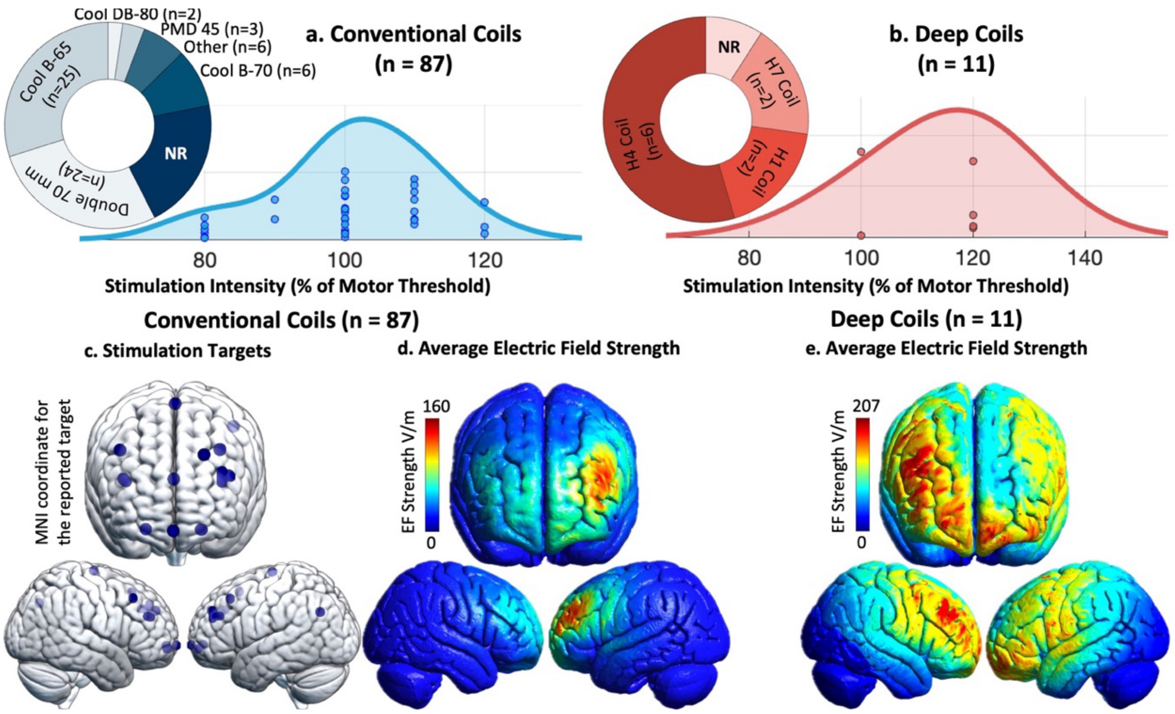


**Figure S3. Visualization of stimulation parameters, targets, and modeled electric fields across conventional and deep TMS protocols.** The upper panels show pie charts summarizing stimulation hardware categories for conventional coils (a) and deep coils (b). Distribution curves depict the spread of stimulation intensities across studies, with individual study values represented as jittered dots. C. presents anatomical maps of reported stimulation targets for conventional TMS coils only, shown as spherical markers plotted onto a standardized cortical surface from superior and bilateral lateral viewpoints. Each dot represents a reported cortical coordinate corresponding to a focal stimulation site used in conventional coil applications. Stimulation targets are not shown for deep coil configurations, as these coils are not designed to stimulate a single coordinate. d and e display three-dimensional cortical renderings of group-averaged modeled electric field distributions for conventional coil protocols (d) and deep coil protocols (e), respectively. Multiple cortical orientations are shown to visualize spatial coverage across hemispheres. For each simulation, coil type, coil placement location, coil orientation, and stimulation intensity were defined based on the protocol parameters reported in the corresponding study to ensure that modeled electric fields reflected study-specific stimulation configurations.

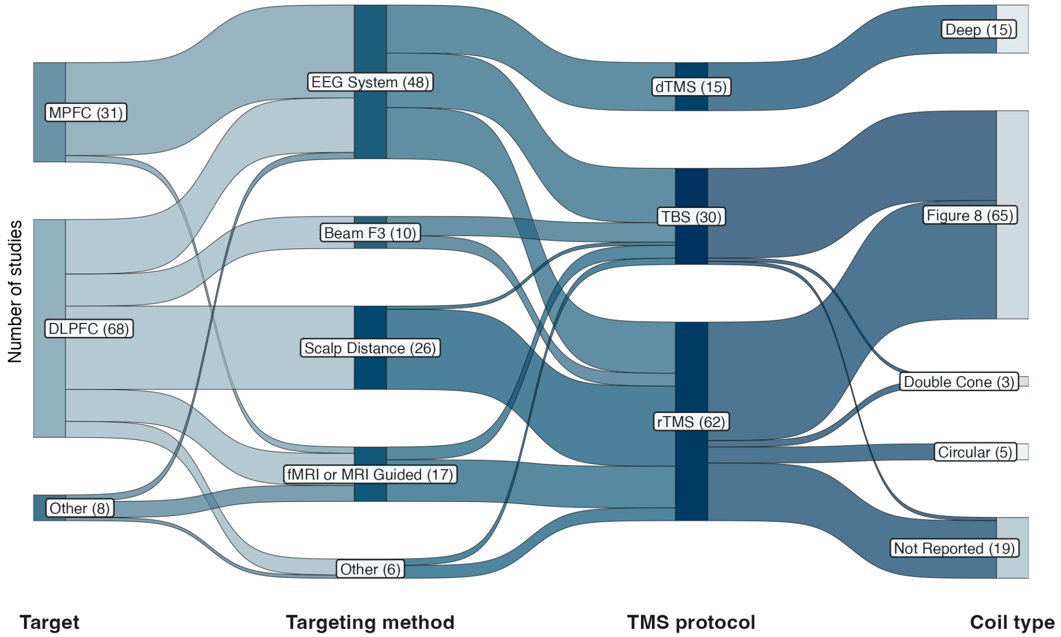


**Figure S4. Variations in target selection and method are visualized.**

**S15. Correlation Map without E-field Normalization and Dividing Conventional and Deep Coils**

In conventional coil analysis, node-wise correlation results showed values ranging from −0.289 to 0.285 (mean = −0.035, SD = 0.104). The 99.9th percentile of positive correlations was r = 0.232, with the peak positive correlation of r = 0.285 localized to the left lateral frontal cortex (MNI −56.4, −17.8, 32.8). The most negative correlation was r = −0.290, centered in the medial frontal cortex (MNI −7.1, 13.4, 9.2).

After permutation testing, five significant clusters were identified: four positive correlation clusters located in the left lateral occipital/posterior parietal cortex (peak at [−49, −57, 32]), left superior parietal cortex/precuneus ([−33, −40, 46]), left ventrolateral prefrontal–orbitofrontal cortex ([−45, 20–22, −16]), and the left fronto-insular region ([−51, 9, 11]), together with a single negative PEC cluster in the right medial prefrontal cortex centered at [18, 25, 6]. In the deep-coil volumetric analysis, tetrahedral node-wise PEC correlations ranged from −0.088 to 0.214 (mean r = 0.099, SD = 0.076). The largest positive correlations reached r = 0.214, with peak values localized to the superior medial frontal/paracentral region (MNI [3.0, 8.2, 51.7]), the midline parietal/precuneus region (MNI [−3.7, −1.5, 28.8]), and an inferior posterior cerebellar/occipital region (MNI [−9.6, −45.5, −43.3]). However, the permutation test showed no significant clusters.

To develop symptom-specific maps, we developed two separate maps for craving and consumption. For the consumption, ranged from r = −0.658 to r = 0.549 (mean = −0.219, SD = 0.186). The strongest negative correlations were localized to the right superior parietal/precuneus region, with peak values at MNI [29.2, −36.1, 58.3] (r = −0.6577), [29.2, −36.4, 59.0] (r = −0.6570), and [31.9, −25.9, 55.4] (r = −0.6549). The highest positive correlations were observed in the ventromedial and orbitofrontal cortex, peaking at MNI [−1.2, 61.7, −17.9] (r = 0.5485), with closely neighboring maxima at [−9.5, 69.6, −5.6] (r = 0.5374) and [−7.1, 69.9, −10.4] (r = 0.5366).

Both corresponding clusters survived significant levels. For the craving map, node-wise correlations ranged from r = −0.245 to r = 0.343 (mean = 0.014, SD = 0.120). The strongest negative correlations clustered within the medial frontal cortex / rostral anterior cingulate region, peaking at MNI [−6.7, 10.6, 9.5] (r = −0.2445) and extending to nearby foci at [−8.2, 11.9, 10.4] (r = −0.2431) and [−7.1, 12.0, 9.7] (r = −0.2429). In contrast, the highest positive correlations were localized to the left lateral frontal cortex (dorsolateral prefrontal cortex region), with peak values at MNI [−56.7, −18.0, 34.6] (r = 0.3428) and adjacent maxima at [−56.4, −18.0, 34.6] (r = 0.3426) and [−55.2, −18.3, 33.3] (r = 0.3426).

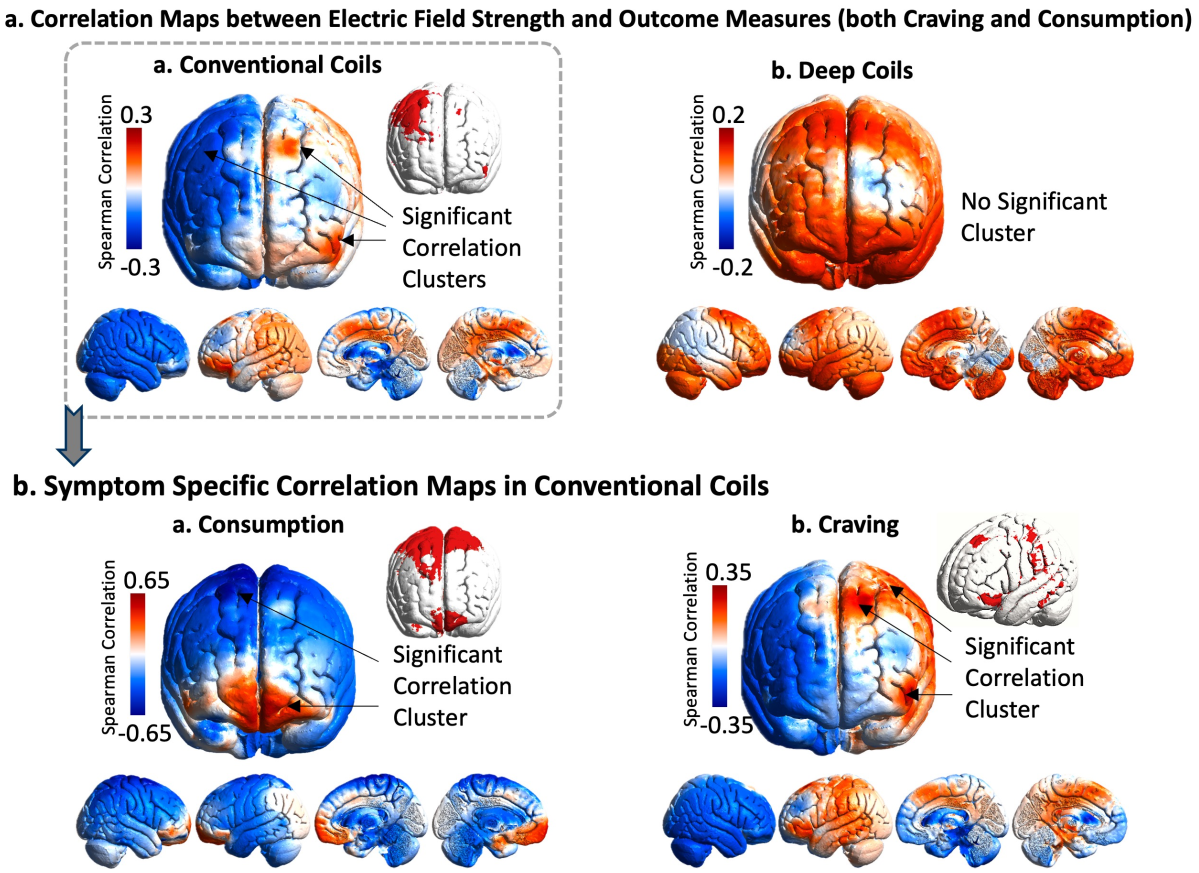


**Figure S5. Electric field–outcome correlation maps for transcranial magnetic stimulation across substance use disorder studies.** (a) Whole-brain node-wise correlation maps between simulated cortical electric field (E-field) strength and clinical effect sizes (Hedges' *g*) for combined craving and consumption outcomes. Spearman correlations were computed across studies for each cortical node using anatomically normalized E-field maps. (a1) Results for conventional coils (not deep) demonstrate spatially localized clusters of significant correlation, indicating cortical regions where higher E-field strength was consistently associated with greater symptom improvement across studies. (a2) Corresponding analyses for deep coils yielded diffuse, non-focal E-field distributions and did not produce statistically significant cortical clusters following permutation-based multiple-comparison correction. (b) Symptom-specific correlation maps restricted to studies using conventional coils. (b1) Consumption outcomes and (b2) craving outcomes were analyzed separately, revealing partially overlapping but distinct cortical clusters exhibiting significant E-field–behavior associations. For all panels, color scales depict Spearman correlation coefficients (warm colors represent positive correlations; cool colors represent negative correlations). Insets show thresholded cortical surfaces highlighting clusters surviving permutation testing. Multiple cortical views are provided to facilitate anatomical interpretation.

#### S16. Replication of the results for one sample standard subject, "Ernie."

To ensure replicability in a standard subject outside MNI space, we repeated the entire pipeline in a healthy standard subject from the SimNIBS sample dataset (Ernie), which has also been used in previous meta-modeling studies (e.g., (24)). Figure S7 illustrates the results for this sample.

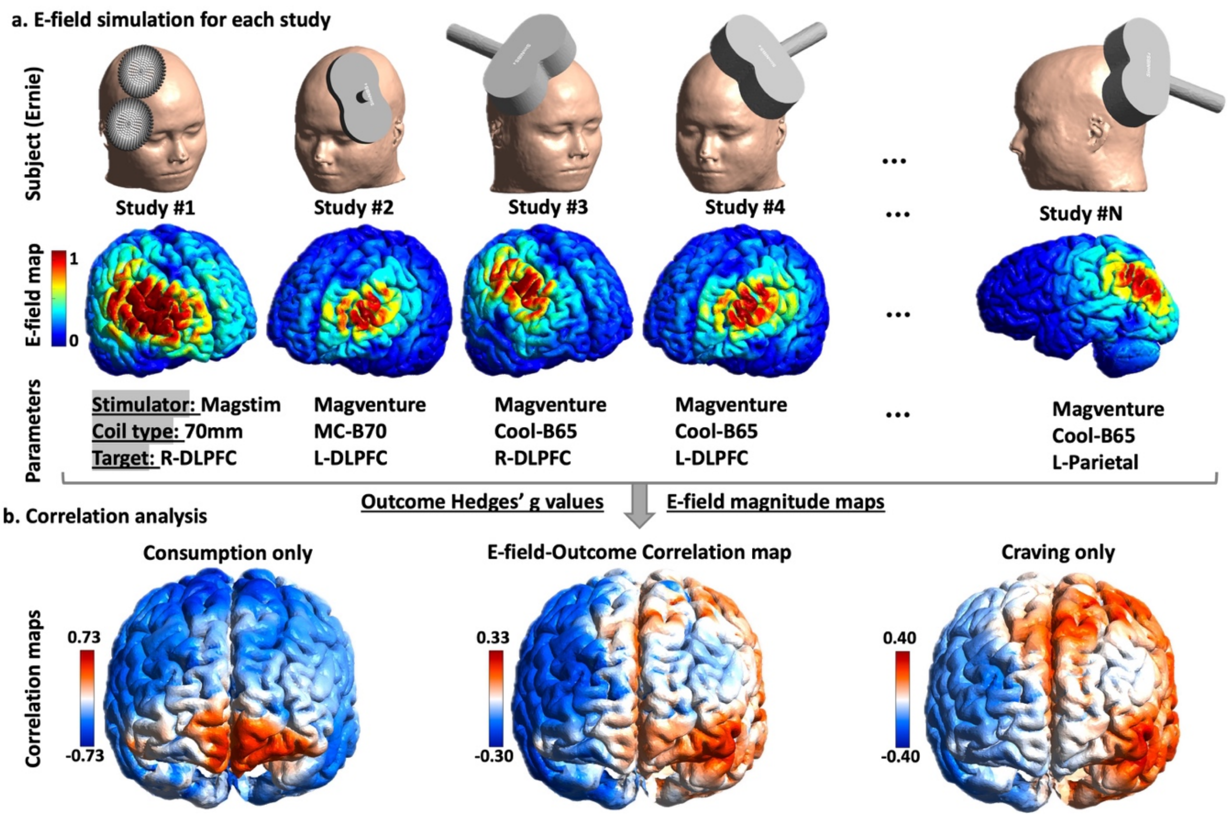


**Figure S6. Visualization of pipeline results in the standard SimNIBS sample subject ("Ernie").** All steps of the pipeline were replicated in the standard head model "Ernie" from the SimNIBS example dataset to demonstrate reproducibility outside MNI space. (a) Illustration of study-specific variability in stimulation parameters, including coil type, stimulator model, and stimulation target, across representative studies. The first row shows the head model with coil placement for each study, the second row shows the corresponding electric field (E-field) magnitude maps, and the third row summarizes the relevant stimulation parameters. (b) Correlation analysis results between voxel-wise E-field magnitude and study-level effect sizes (Hedges' g). The middle map shows correlations across both craving and consumption outcomes combined, the right map shows craving-only analyses, and the left map shows consumption-only analyses.

#### S17. Group-level results for the clinical population

Individualized computational head models were constructed for all 60 participants with methamphetamine use disorder (MUD). The full correlation-mapping pipeline was executed independently for each participant. Subject-specific maps were then projected onto the *fsaverage* surface to harmonize spatial resolution and topology, after which population-level mean correlation maps were derived (Figure S6).

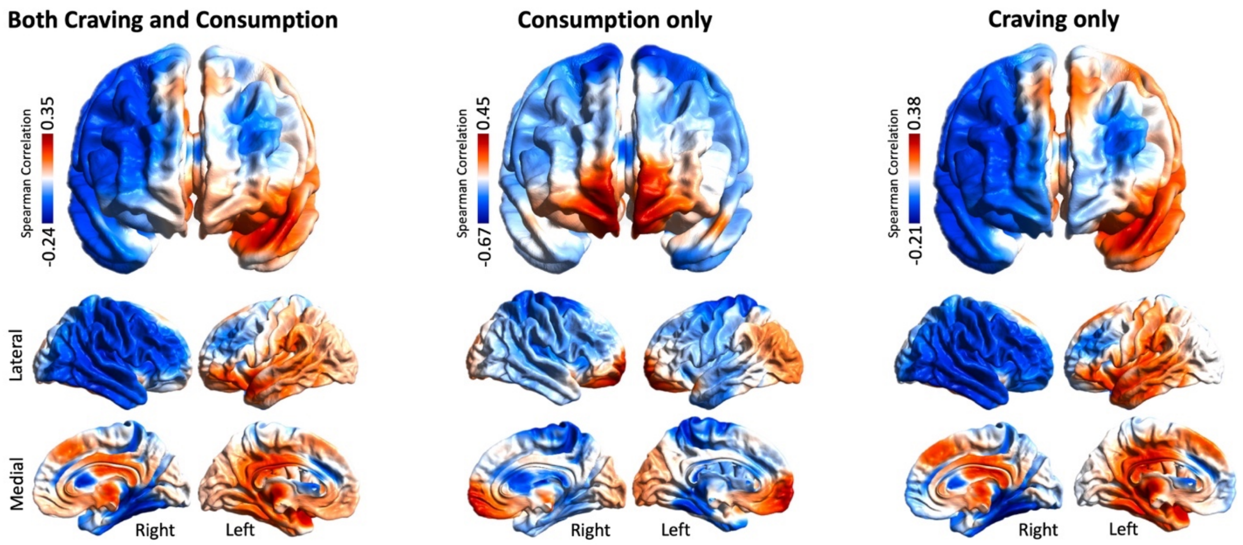


**Figure S7. Unthresholded electric field–outcome correlation maps on the fsaverage cortical surface for 60 participants with methamphetamine use disorders.** Whole-brain, node-wise Spearman correlation maps illustrate the relationship between simulated cortical electric field (E-field) strength and clinical effect sizes (Hedges' *g*) across transcranial magnetic stimulation studies. Maps are shown separately for outcomes reflecting **both craving and consumption** (left), **consumption only** (middle), and **craving only** (right). Correlations were computed across studies at each cortical node using anatomically normalized E-field maps and projected onto the *fsaverage* surface to ensure a common spatial framework. Lateral and medial views of both hemispheres are displayed to facilitate anatomical interpretation. Color scales represent Spearman correlation coefficients, with warm colors indicating positive associations and cool colors indicating negative associations.

#### S18. Protocol-Specific E-field outcome specific

To address the potential influence of stimulation protocol on the observed E-field–outcome associations, we conducted additional exploratory analyses stratified by protocol category. Studies were grouped into two categories: (1) high-frequency rTMS and intermittent theta burst stimulation (HF-rTMS/iTBS), and (2) low-frequency rTMS and continuous theta burst stimulation (LF-rTMS/cTBS). Protocol-specific E-field–effect size association maps were generated using the same analytical framework described in the main manuscript. 70.4% were classified as HF-rTMS/iTBS, other as LF-rTMS/cTBS. Because the number of LF-rTMS/cTBS and dTMS studies was substantially smaller than the number of HF-rTMS/iTBS studies, these analyses should be interpreted as exploratory and descriptive rather than confirmatory.

Supplementary Figure S9 presents the protocol-specific E-field–effect size association maps. Broadly, the spatial patterns observed for HF-rTMS/iTBS were similar to those identified in the primary analysis, reflecting the fact that these protocols constituted the majority of the available literature. Although LF-rTMS/cTBS maps showed partially overlapping patterns, differences in the spatial distribution and directionality of associations were observed. However, the limited number of available studies in the LF-rTMS/cTBS and dTMS categories precluded formal statistical comparisons between protocol groups and limited the ability to draw definitive conclusions regarding protocol-specific effects.

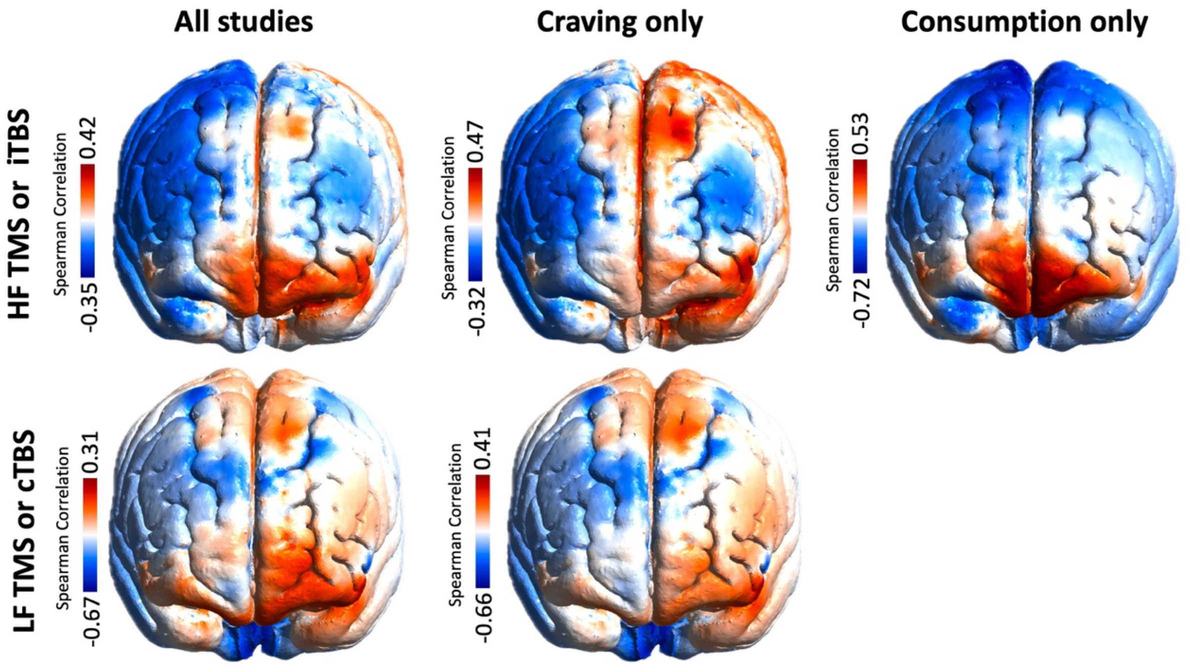


**Supplementary Figure S8. Protocol-specific E-field–effect size association maps.** Voxel-wise Spearman correlations between simulated E-field magnitude and treatment effect size are shown separately for high-frequency rTMS/intermittent theta burst stimulation (HF-rTMS/iTBS; top row) and low-frequency rTMS/continuous theta burst stimulation (LF-rTMS/cTBS; bottom row). Correlation maps are displayed for all outcome measures combined (left column), craving outcomes only (middle column), and consumption outcomes only (right column). Positive correlations (red-yellow) indicate regions where stronger induced E-fields were associated with larger treatment effects, whereas negative correlations (blue) indicate the opposite relationship.

To further address the possibility that stimulation protocol influenced the observed E-field–outcome associations, we conducted an additional voxel-wise sensitivity analysis restricted to HF-rTMS/iTBS ($n = 69$) and LF-rTMS/cTBS ($n = 18$) studies. At each mesh element, treatment effect size was modeled as a function of local E-field magnitude, protocol category, and an E-field × protocol interaction term:

$$\text{Hedges' } g = \beta_0 + \beta_1(\text{E-field}) + \beta_2(\text{Protocol}) + \beta_3(\text{E-field} \times \text{Protocol}) + \varepsilon$$

where Protocol was coded as HF-rTMS/iTBS (0) or LF-rTMS/cTBS (1). The $\beta_1$ coefficient was used to generate protocol-adjusted E-field association maps, whereas $\beta_3$ quantified the E-field × Protocol interaction. The protocol-adjusted E-field association maps were broadly similar to those observed in the primary analysis, indicating that the principal spatial relationships between induced E-field magnitude and treatment outcome remained evident after accounting for stimulation protocol. Examination of the E-field × Protocol interaction maps revealed no robust or spatially consistent interaction effects. Interaction statistics remained low across most cortical regions and did not identify large clusters suggestive of protocol-dependent E-field–outcome relationships. These findings suggest that the primary E-field–outcome associations were not solely driven by differences between HF-rTMS/iTBS and LF-rTMS/cTBS protocols and support the stability of the main results across stimulation paradigms.

#### S19. Left DLPFC–Restricted E-Field–Outcome Association Mapping

To evaluate whether the spatial associations identified in the primary analysis were driven by heterogeneity in stimulation target location, we performed an additional sensitivity analysis restricted to studies targeting the left dorsolateral prefrontal cortex (DLPFC). Studies were included if the stimulation target was classified as left DLPFC based on the targeting approach reported in the original publication (e.g., F3, Beam F3, 5-cm rule, MRI-guided left DLPFC targeting, or equivalent left prefrontal targets).

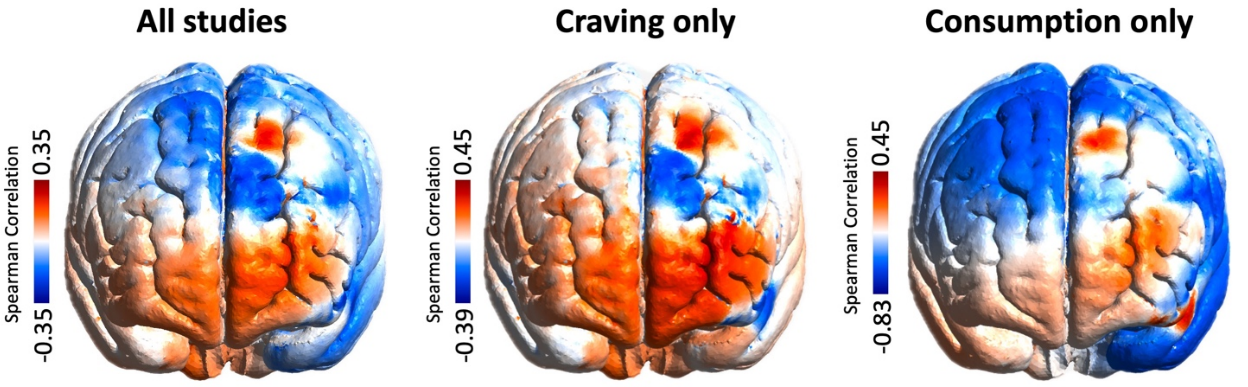


**Supplementary Figure S9. Left DLPFC-restricted E-field–outcome association maps.** Element-wise Spearman correlation coefficients between normalized electric-field (E-field) magnitude and study-level treatment effects (Hedges' g) across studies targeting the left dorsolateral prefrontal cortex (DLPFC). Analyses were performed using all outcomes combined (left), craving outcomes only (middle), and consumption outcomes only (right). Warm colors indicate cortical regions where greater induced E-field magnitude was associated with larger treatment effects across studies, whereas cool colors indicate regions where greater E-field magnitude was associated with smaller treatment effects. Correlation patterns were broadly consistent across analyses, with positive associations concentrated in ventral prefrontal, orbitofrontal, and frontopolar regions and negative associations observed primarily in dorsal frontal and parietal areas.

#### S20. Stability of the correlation maps

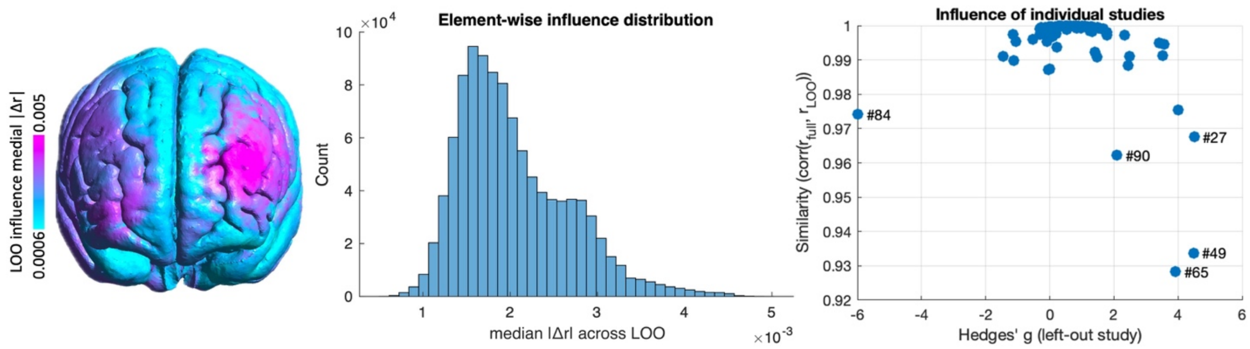


**Figure S10. Leave-one-out (LOO) influence and robustness of the correlation map. Left:** Cortical surface visualization of the element-wise LOO influence map, defined as the median absolute change in correlation values ($|\Delta r|$) across all LOO iterations. Warmer colors indicate regions where E-field-behavior correlation values are more sensitive to the exclusion of individual studies, whereas cooler colors reflect highly stable regions. Overall influence values were small across the cortex, indicating strong spatial robustness of the E-field-behavior correlation map. **Middle:** Histogram of element-wise LOO influence values (median $|\Delta r|$ across iterations). The distribution is right-skewed, with the majority of elements exhibiting minimal influence, demonstrating that most regions show negligible variability when individual studies are removed. **Right:** Scatter plot showing the relationship between the effect size (Hedges' *g*) of the left-out study and the similarity between the full E-field-behavior map and the corresponding LOO map, quantified as Pearson correlation. Each point represents one LOO iteration, with selected influential iterations labeled. High similarity values across a wide range of effect sizes indicate that no single study disproportionately drives the overall E-field-behavior correlation map spatial pattern.

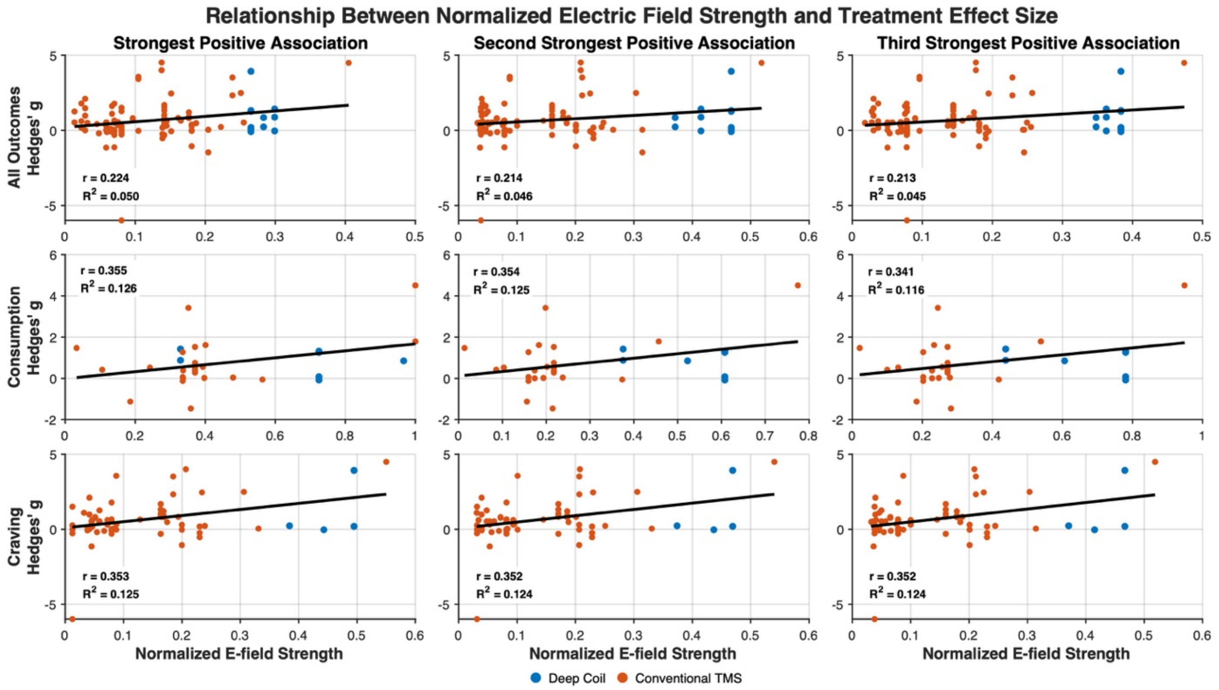


**Figure S11. Relationship between normalized electric field strength and clinical effect size across TMS studies.** Scatter plots illustrate the association between normalized electric field (E-field) strength (x-axis) and treatment effect size (Hedges' *g*; y-axis) across included TMS studies. Rows correspond to analyses including all outcomes (top row), consumption outcomes only (middle row), and craving outcomes only (bottom row). Columns display the three cortical nodes exhibiting the strongest positive E-field–effect size associations within each correlation map. Each point represents an individual study outcome measure. Blue markers indicate deep TMS coils, whereas orange markers indicate conventional TMS coils. E-field values were normalized to account for systematic differences in field strength across coil types. Solid black lines represent linear regression fits across all studies included in each analysis. The corresponding Pearson correlation coefficient (*r*) and coefficient of determination ($R^2$) are shown within each panel. Overall, positive associations between local E-field strength and treatment effect size were observed across the identified nodes, with stronger associations evident for craving and consumption outcomes than for the combined analysis.

#### What do we have over DLPFC?

The left DLPFC was the most frequently targeted brain region across TMS studies; however, no significant correlation was observed between E-field strength in the left DLPFC and effect sizes. Although the estimated correlation was slightly negative ($r < -0.1$), the scatter plot reveals substantial variability in effect sizes at comparable normalized E-field levels.

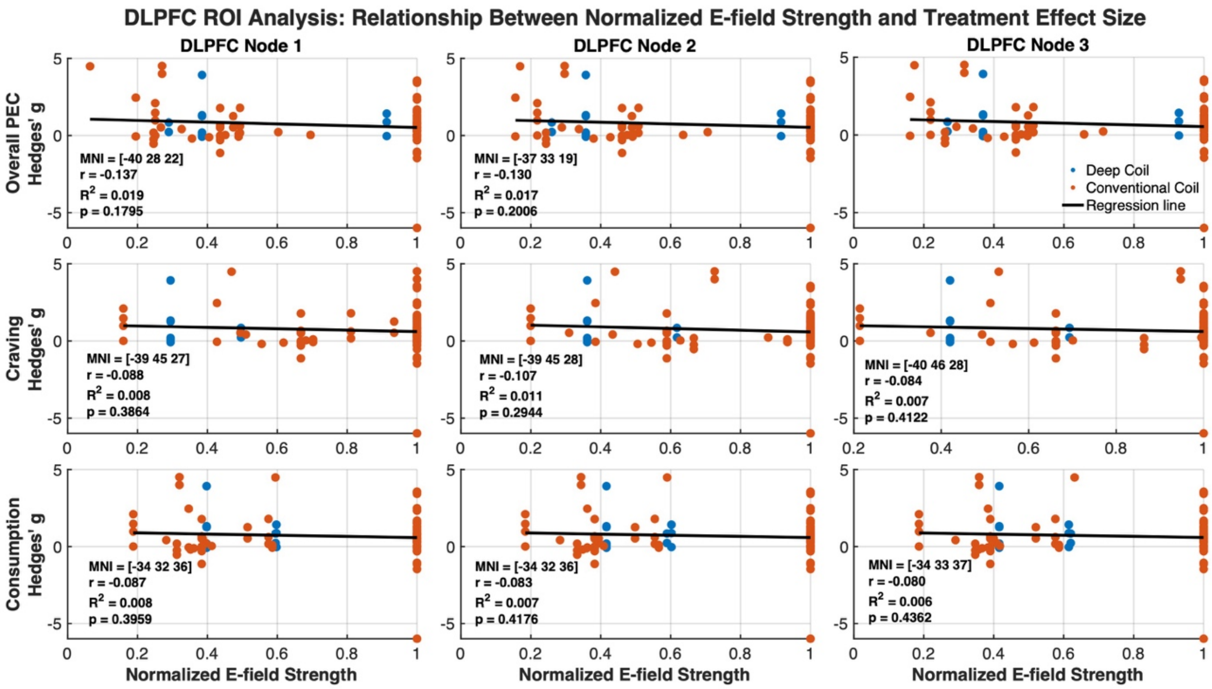


**Figure S12. Relationship between normalized electric field strength and clinical effect size across TMS studies within the left DLPFC.** Scatter plots illustrating the association between normalized electric field strength (95th-percentile normalized and capped; p95cap) and treatment effect size (Hedges' *g*) at selected nodes within a predefined left DLPFC region of interest (15-mm radius sphere centered at MNI [−44, 40, 29]). Rows correspond to the overall E-field–effect size association map (top row), craving-specific analysis (middle row), and consumption-specific analysis (bottom row). Within each row, the three columns represent the nodes exhibiting the strongest negative E-field–effect size associations for that specific analysis. Each point represents an individual study. Blue markers indicate Deep Coil TMS protocols, whereas orange markers indicate Conventional TMS protocols. Solid black lines denote least-squares linear regression fits. For each panel, the corresponding MNI coordinates, Pearson correlation coefficient ($r$), coefficient of determination ($R^2$), and associated $p$-value are displayed. Normalized E-field values were derived using study-wise 95th-percentile normalization to facilitate comparison across stimulation protocols with different stimulation intensities and coil configurations.

Specifically, many studies cluster at similar high normalized E-field values, yet report widely divergent Hedges' g values ranging from strongly negative to strongly positive. This pattern indicates that similar local E-field strength in the left DLPFC can be associated with markedly different clinical effects. Conversely, several studies with lower E-field strength in the left DLPFC—often due to targeting other regions such as the right DLPFC—reported relatively larger effect sizes. Together, these observations suggest that factors beyond local E-field magnitude, including study population characteristics, stimulation protocol parameters (e.g., number of sessions), and baseline neural state, substantially contribute to outcome variability. As a result, the modest negative trend observed over the left DLPFC does not reach statistical significance, reflecting heterogeneity in study design and biological context rather than the absence of a neuromodulatory effect.

This pattern reflects a methodological limitation of correlation-based mapping in regions with restricted E-field variance rather than evidence against the relevance of the left DLPFC as a neuromodulation target. Because many studies delivered similar levels of stimulation to the left

DLPFC, normalized E-field values in this region show limited variability across studies. In contrast, effect sizes vary widely due to differences in study populations, stimulation protocols, and outcome measures. Under these conditions of range restriction, correlation analyses have limited power to detect meaningful associations, even when the targeted region is functionally relevant. Consequently, the observed weak and non-significant correlation indicates that local E-field magnitude alone does not account for outcome variability across studies, rather implies a lack of therapeutic relevance of left DLPFC stimulation.

**S21. K-fold Robustness Analysis**

To further assess the robustness of the E-field–outcome association maps, we performed a K-fold resampling analysis. For each analysis, studies were randomly partitioned into either 5 or 10 approximately equal folds. The E-field–outcome association map was first computed using the full dataset, following the same procedure described in the main manuscript. Subsequently, for each fold, all studies belonging to that fold were excluded, and the association map was recomputed using the remaining studies. Spatial similarity between each fold-derived map and the full-sample map was quantified across all gray-matter elements using both Pearson and Spearman correlations. This procedure was repeated across multiple random fold assignments, generating a distribution of similarity metrics for each partition scheme (5-fold and 10-fold). High similarity between fold-derived maps and the full-sample map would indicate that the observed spatial associations are not driven by a small subset of studies and are robust to the exclusion of substantial portions of the dataset.

The spatial distribution of E-field–outcome associations remained highly stable across K-fold resampling analyses. For the 5-fold partition, the mean Pearson correlation between fold-derived maps and the full-sample map was 0.94 ± 0.05, while the mean Spearman correlation was 0.94 ± 0.05. Similarly, for the 10-fold partition, the mean Pearson correlation was 0.97 ± 0.03 and the mean Spearman correlation was 0.97 ± 0.03. These results demonstrate that the overall spatial pattern of E-field–outcome associations was highly reproducible even when substantial subsets of studies were excluded from the analysis. The slightly higher similarity observed in the 10-fold analysis is expected because a smaller proportion of studies is removed in each iteration. Importantly, both partition schemes produced nearly identical spatial maps, indicating that the identified associations were not driven by individual studies or small subsets of the dataset.

**S22. Expanded Discussion of Study Limitations and Future Directions**

Despite the robustness of our findings, several limitations should be considered. (1) Heterogeneity in outcome assessment, including differences in measurement instruments and timing for craving and consumption, represents an important source of variability across studies, despite efforts to harmonize outcomes.

(2) The present framework is based on study-level associations; the E-field–effect size maps reflect statistical relationships rather than direct causal effects, even though non-invasive brain stimulation is itself a causal intervention (21). Within this approach, the ability to detect E-field–behavior relationships depends on sufficient variability in stimulation parameters across studies (see Supplementary S5 for an example using DLPFC targeting). Although such variability was

present in the current dataset and enabled robust spatial associations, it may limit generalizability to more homogeneous stimulation protocols in other neuropsychiatric contexts. The main point about this approach is that meta-modeling is inherently more sensitive to identifying E-field–outcome associations in regions that exhibit substantial between-study variability in induced field strength. Consequently, regions such as the frontopolar cortex, IFG, and pre-SMA may emerge not only because they are functionally relevant to addiction, but also because they received more variable incidental E-field exposure across studies. In contrast, regions that were consistently stimulated with similar E-field intensities across studies may show weaker associations due to restricted variability, regardless of their potential clinical importance. Therefore, the identified regions should be interpreted as locations where variability in E-field engagement was associated with variability in treatment outcomes, rather than as definitive evidence of therapeutic superiority or optimal stimulation targets.

(3) The spatial overlap analysis between the FDCR activation map and the E-field–effect size association map should be interpreted as supportive evidence rather than as a validation of the broader meta-modeling framework. Our overlap analysis was conducted in a single cohort of individuals with MUD, whereas the meta-analytic dataset included multiple SUD populations and was predominantly composed of nicotine- and alcohol-related studies. Therefore, the generalizability of these findings across substance types remains uncertain and should be evaluated in future studies using independent cohorts and other SUD populations. Additionally, significant spatial overlap was observed only for the consumption-related association map and not for the craving-related association map. Because drug cue reactivity paradigms are conceptually more proximal to craving than to substance consumption, the interpretation of this overlap would be complicated. One possible explanation is that cue-reactivity–related neural responses capture broader motivational and behavioral processes that contribute to downstream substance use behavior. Nevertheless, the observed overlap should be interpreted as evidence of spatial correspondence rather than validation of consumption-specific mechanisms or of the meta-modeling framework itself.

(4) Although the present E-field modeling framework incorporated stimulation intensity by scaling simulated electric fields according to the reported stimulation intensity (primarily percentage of resting motor threshold) and device-specific dI/dt values, stimulation intensity represents only one component of the overall TMS dose. Additional parameters, including the total number of pulses, number of sessions, sessions per day, treatment duration, and inter-session intervals, may also influence treatment outcomes and contribute to variability in observed effect sizes. These dosing parameters operate at multiple temporal scales, including within-session stimulation characteristics (e.g., frequency, burst structure, train duration, and pulse number) and between-session treatment schedules (e.g., number and spacing of sessions). Consequently, two protocols with similar instantaneous E-field strengths may differ substantially in their cumulative neurobiological effects. Furthermore, repeated-session and accelerated stimulation protocols may engage distinct mechanisms of neural plasticity even when cumulative stimulation exposure is comparable. The current analysis did not explicitly model potential nonlinear dose–response relationships or interactions among multiple dosing parameters. Future studies incorporating detailed stimulation dose metrics and longitudinal treatment schedules

may help clarify how E-field characteristics interact with temporal aspects of TMS dosing to influence clinical outcomes.

(5) Only a limited number of addiction studies included in the present dataset used fMRI-guided target definition, and even in those studies, the reported stimulation targets were represented as group-level coordinates rather than participant-specific target locations. Consequently, the current meta-modeling framework was restricted to study-level target definitions and could not evaluate variability arising from individualized target selection. However, the use of participant-specific fMRI-guided targeting is becoming increasingly common in TMS research. As these data become more widely available, future meta-modeling studies could incorporate individualized stimulation coordinates even across a specific anatomical region (e.g., left DLPFC based on the most negative correlation with sgACC for each participant), head models, and behavioral outcomes. Such datasets would enable E-field–outcome association maps to be generated at the participant level rather than the study level, potentially providing a more precise characterization of how variability in target location and E-field engagement relates to clinical outcomes.

Finally, consistent with prior studies using similar meta-modeling approaches (21, 24, 25, 26), the present framework relies on study-level effect sizes rather than individual participant data. This limits the ability to model within-study variability, individual-level dose–response relationships, and participant-specific factors such as symptom severity or abstinence duration. Although we partially addressed inter-individual variability by replicating the pipeline in a clinical sample using anatomically individualized head models, the broader literature still lacks studies that integrate individual MRI data, detailed stimulation parameters, and longitudinal behavioral outcomes. This limits the extent to which individual-level inferences can be drawn. As a future direction, large-scale data-sharing initiatives such as the ENIGMA–Neuromodulation working group (27) may facilitate access to individual-level neuroimaging data, enabling more accurate E-field simulations and next-generation meta-modeling approaches.

#### Craving

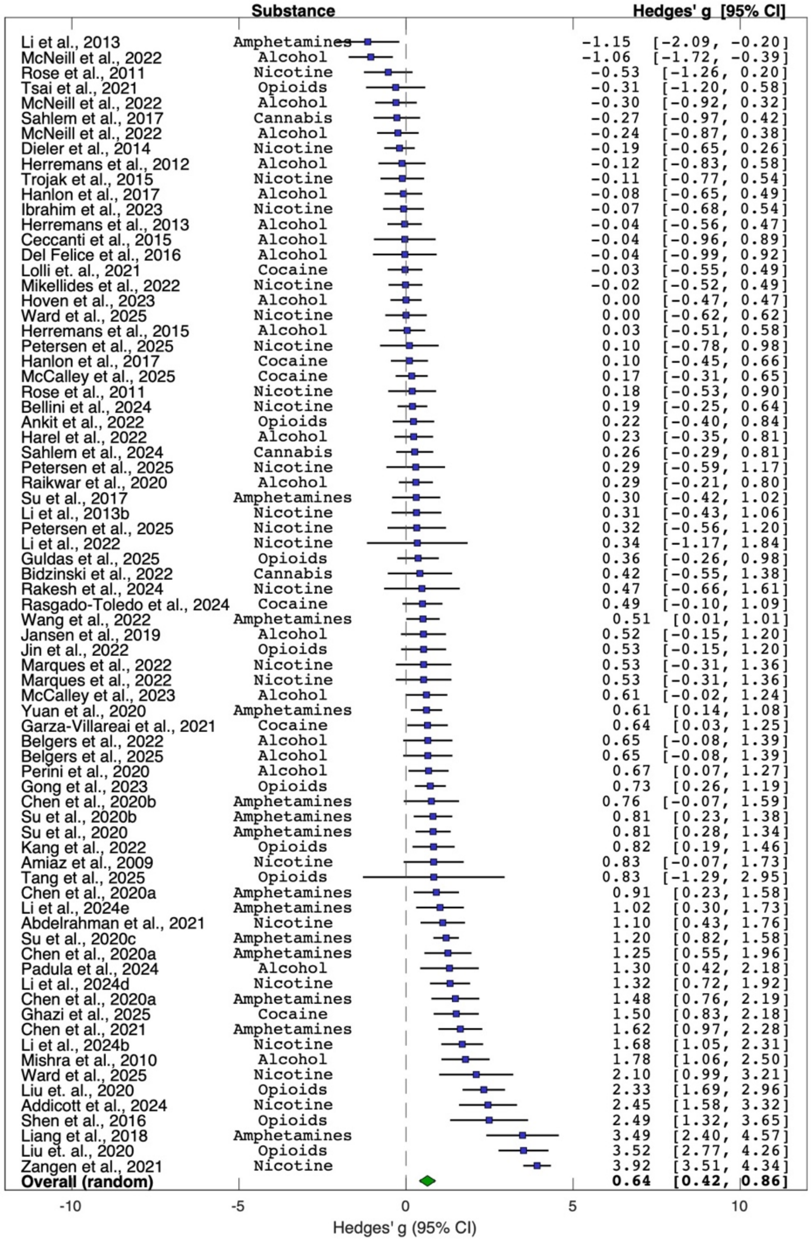


#### Consumption

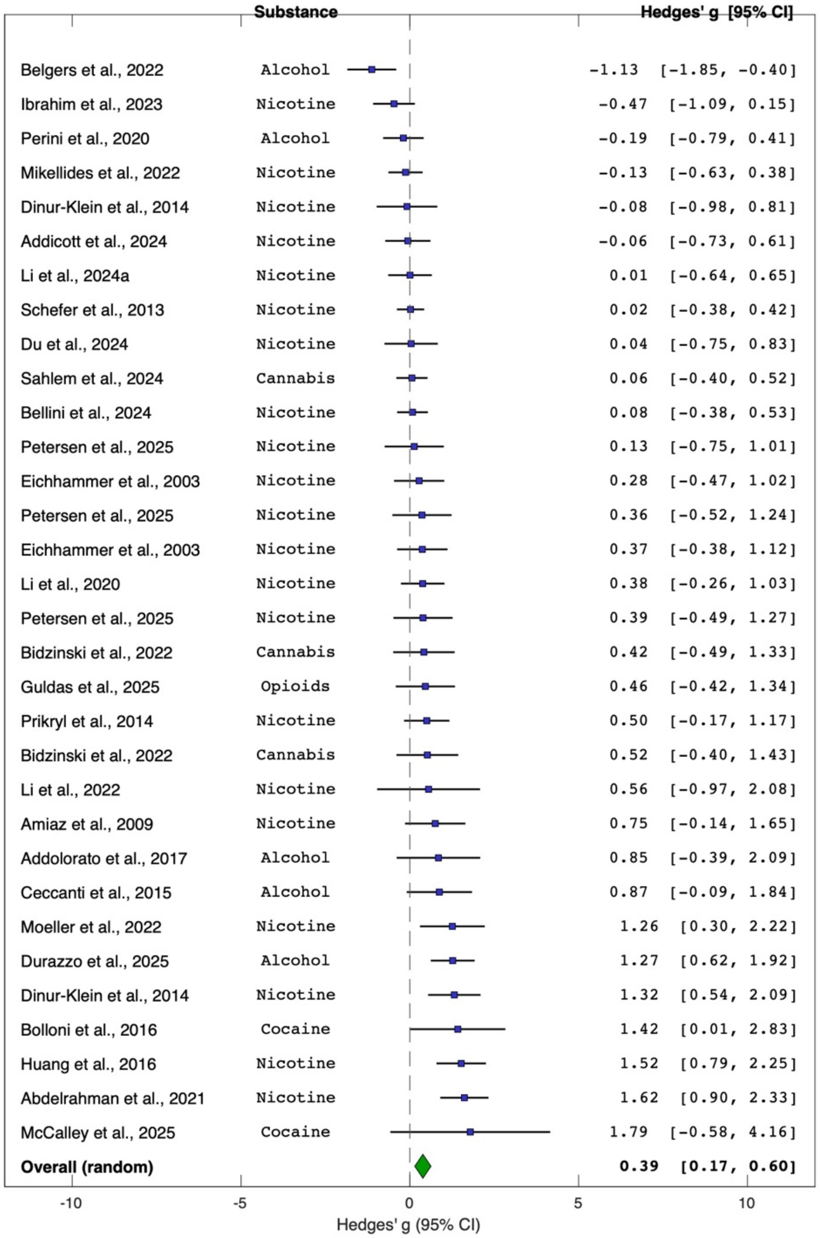

#### Table S1. Data extraction from included TMS studies that reported craving or consumption as an outcome measure

| Study (Author and Year) | Ref | Addiction type | Sham subjects | Active subjects | Targeting method | Intensity RMT% | Sessions | TMS type | Frequency | Pulses/session | Coil type | Time point | Scale unit | Outcome measure | Hedges' g |
|---|---|---|---|---|---|---|---|---|---|---|---|---|---|---|---|
| (Eichhammer et al., 2003) | (5) | Nicotine | 14 | 14 | 5 cm rule | 90 | 2 | rTMS | 20 | 1000 | Magstim_70mm_Fig8 | T0 | Self-reported (cigarettes per day or hour) | Consumption | 0.37 |
| (Eichhammer et al., 2003) | (5) | Nicotine | 14 | 14 | 5 cm rule | 90 | 2 | rTMS | 20 | 1000 | Magstim_70mm_Fig8 | T0 | Self-reported (cigarettes per day or hour) | Consumption | 0.28 |
| (Amiaz et al., 2009) | (6) | Nicotine | 9 | 12 | 5 cm rule | 100 | 10 | rTMS | 10 | 1000 | Magstim_70mm_Fig8 | T0 | Self-reported (cigarettes per day or hour) | Consumption | 0.75 |
| (Sheffer et al., 2013) | (28) | Nicotine | 47 | 47 | 6 cm rule | 110 | 1 | rTMS | 20/ 10 | 900 | Magstim_70mm_Fig8 | Follow Up | Self-reported (cigarettes per day or hour) | Consumption | 0.02 |
| (Dinur-Klein et al., 2014) | (29) | Nicotine | 15 | 7 | Fz | 120 | 13 | dTMS | 10 | 900 | H4 | T0 | Self-reported (cigarettes per day or hour) | Consumption | -0.08 |
| (Dinur-Klein et al., 2014) | (29) | Nicotine | 15 | 16 | Fz | 120 | 13 | dTMS | 1 | 600 | H4 | T0 | Self-reported (cigarettes per day or hour) | Consumption | 1.32 |
| (Prikryl et al., 2014) | (30) | Nicotine | 17 | 18 | 5 cm rule | 110 | 21 | rTMS | 10 | 2000 | Magstim_70mm_Fig8 | T0 | Self-reported (cigarettes per day or hour) | Consumption | 0.50 |
| (Ceccanti et al., 2015) | (31) | Alcohol | 9 | 9 | F1 | 120 | 10 | dTMS | 20 | 1500 | H1 | T0 | Self-reported (alcoholic drinks per day) | Consumption | 0.87 |
| (Bolloni et al., 2016) | (32) | Cocaine | 4 | 6 | F1 | 100 | 12 | dTMS | 10 | 1000 | H1 | T0 | Unitary cocaine amount | Consumption | 1.42 |
| (Huang et al., 2016) | (33) | Nicotine | 18 | 19 | 5 cm rule | 110 | 21 | rTMS | 10 | 2000 | Magstim_70mm_Fig8 | T0 | Self-reported (cigarettes per day or hour) | Consumption | 1.52 |
| (Addolorato et al., 2017) | (34) | Alcohol | 6 | 5 | AFz | 100 | 12 | dTMS | 10 | 1000 | H7 | T0 | Timeline Followback (TLFB) | Consumption | 0.85 |
| (Li et al., 2020) | (15) | Nicotine | 17 | 21 | Neuronavigation | 100 | 10 | rTMS | 10 | 3000 | Magstim_70mm_Fig8 | T0 | Self-reported (cigarettes per day or hour) | Consumption | 0.38 |
| (Perini et al., 2020) | (35) | Alcohol | 20 | 23 | AFz | 120 | 15 | dTMS | 10 | 1500 | H8 | T0 | Serum Phosphatidylethanol (PEth) | Consumption | -0.19 |
| (Abdelrahman et al., 2021) | (36) | Nicotine | 20 | 20 | 7 cm rule | 80 | 10 | rTMS | 20 | 2000 | Magstim_70mm_Fig8 | T0 | Self-reported (cigarettes per day or hour) | Consumption | 1.62 |
| (Belgers et al., 2022) | (37) | Alcohol | 18 | 16 | F4 | 110 | 10 | rTMS | 10 | 3000 | Magstim_70mm_Fig8 | Follow Up | Timeline Followback (TLFB) | Consumption | -1.13 |
| (Bidzinski et al., 2022) | (4) | Cannabis | 10 | 9 | F5 | 90 | 20 | rTMS | 20 | 750 | MagVenture_Cool-B65 | T0 | Self-reported (substance use) | Consumption | 0.52 |
| (Bidzinski et al., 2022) | (4) | Cannabis | 10 | 9 | F6 | 90 | 20 | rTMS | 20 | 750 | MagVenture_Cool-B65 | T0 | Self-reported (substance use) | Consumption | 0.42 |
| (Mikellides et al., 2022) | (38) | Nicotine | 30 | 30 | Beam F3 | 100 | 20 | iTBS | 5 | 600 | MagVenture_Cool-B65 | T0 | Self-reported (cigarettes per day or hour) | Consumption | -0.13 |
| (Moeller et al., 2022) | (39) | Nicotine | 10 | 10 | Fz | 120 | 15 | dTMS | 10 | 1800 | H4 | T0 | Self-reported (cigarettes per day or hour) | Consumption | 1.26 |
| (Li et al., 2022) | (40) | Nicotine | 4 | 3 | 5.5 cm rule | 100 | 5 | rTMS | 10 | 3000 | Magstim_70mm_Fig8 | T0 | Self-reported (cigarettes per day or hour) | Consumption | 0.56 |
| (Ibrahim et al., 2023) | (41) | Nicotine | 18 | 24 | NA | 120 | 20 | dTMS | 10 | 1020 | H11 | T0 | Timeline Followback (TLFB) | Consumption | -0.47 |

| | | | | | | | | | | | | | | | |
|---|---|---|---|---|---|---|---|---|---|---|---|---|---|---|---|
| **(X. Li et al., 2024)** | **(42)** | Nicotine | 17 | 20 | Neuronavigation | 100 | 10 | rTMS | 10 | 3000 | Magstim_70mm_Fig8 | T0 | Self-reported (cigarettes per day or hour) | Consumption | 0.01 |
| **(Du et al., 2024)** | **(43)** | Nicotine | 10 | 16 | fMRI | 100 | 20 | rTMS | 10 | 1200 | Magstim_70mm_Fig8 | T0 | Self-reported (cigarettes per day or hour) | Consumption | 0.04 |
| **(Sahlem et al., 2024)** | **(11)** | Cannabis | 35 | 37 | Beam F3 | 120 | 20 | rTMS | 10 | 4000 | MagVenture_Cool-B65 | T0 | Self-reported (days per week of Cannabis Use) | Consumption | 0.06 |
| **(Bellini et al., 2024)** | **(44)** | Nicotine | 37 | 37 | Fz | 120 | 21 | dTMS | 10 | 1800 | H4 | T0 | Self-reported (cigarettes per day or hour) | Consumption | 0.08 |
| **(Addicott et al., 2024)** | **(45)** | Nicotine | 13 | 25 | AFz | 110 | 20+8 | iTBS | 5 | 600 | MagVenture_Cool-B65 | T0 | Self-reported (cigarettes per day or hour) | Consumption | -0.06 |
| **(Durazzo et al., 2025)** | **(46)** | Alcohol | 22 | 22 | Beam F3 | 110 | 20 | iTBS | 5 | 1200 | MagVenture_Cool-B65 | Follow Up | Timeline Followback (TLFB) | Consumption | 1.27 |
| **(D. M. McCalley et al., 2025)** | **(47)** | Cocaine | 12 | 13 | Fp1 | 110 | 10 | cTBS | 5 | 1800 | MagVenture_Cool-B65 | Follow Up | Timeline Followback (TLFB) | Consumption | 1.79 |
| **(Petersen et al., 2025)** | **(14)** | Nicotine | 60 | 66 | fMRI | 100 | 1 | rTMS | 10 | 3000 | MagVenture_Cool-B65 | T0 | Shiffman–Jarvik Withdrawal Scale (SJWS) | Consumption | 0.36 |
| **(Petersen et al., 2025)** | **(14)** | Nicotine | 61 | 66 | fMRI | 100 | 1 | rTMS | 10 | 3000 | MagVenture_Cool-B65 | T0 | Shiffman–Jarvik Withdrawal Scale (SJWS) | Consumption | 0.39 |
| **(Petersen et al., 2025)** | **(14)** | Nicotine | 62 | 66 | fMRI | 100 | 1 | rTMS | 10 | 3000 | MagVenture_Cool-B65 | T0 | Shiffman–Jarvik Withdrawal Scale (SJWS) | Consumption | 0.13 |
| **(Guldas et al., 2025)** | **(48)** | Opioids | 18 | 21 | 5 cm rule | 110 | 20 | rTMS | 10 | 3000 | Cool-DB80 | Follow Up | Unitary cocaine amount | Consumption | 0.46 |
| **(Amiaz et al., 2009)** | **(6)** | Nicotine | 9 | 12 | 5 cm rule | 100 | 10 | rTMS | 10 | 1000 | Magstim_70mm_Fig8 | T0 | Visual Analogue Scale (VAS) | Craving | 0.83 |
| **(Mishra et al., 2010)** | **(49)** | Alcohol | 15 | 30 | Not Reported | 110 | 10 | rTMS | 10 | 1000 | Magstim_70mm_Fig8 | T0 | Alcohol Craving Questionnaire (ACQ) | Craving | 1.78 |
| **(Rose et al., 2011)** | **(50)** | Nicotine | 15 | 15 | Fpz | 90 | 1 | rTMS | 10 | 600 | Magstim_70mm_Fig8 | T0 | Shiffman-Jarvik Withdrawal Scale (SJWS) | Craving | -0.53 |
| **(Rose et al., 2011)** | **(50)** | Nicotine | 15 | 15 | Fpz | 90 | 1 | rTMS | 1 | 600 | Magstim_70mm_Fig8 | T0 | Shiffman-Jarvik Withdrawal Scale (SJWS) | Craving | 0.18 |
| **(S. Herremans et al., 2012)** | **(16)** | Alcohol | 16 | 15 | Neuronavigation | 110 | 1 | rTMS | 20 | 1560 | Magstim_70mm_Fig8 | T0 | Obsessive Compulsive Drinking Scale (OCDS) | Craving | -0.12 |
| **(S. Herremans et al., 2013)** | **(51)** | Alcohol | 29 | 29 | F4 | 110 | 1 | rTMS | 20 | 1560 | Magstim_70mm_Fig8 | T0 | Obsessive Compulsive Drinking Scale (OCDS) | Craving | -0.04 |
| **(Li, Malcolm, et al., 2013)** | **(9)** | Meth | 10 | 10 | 6 cm rule | 100 | 1 | rTMS | 1 | 900 | Magstim_70mm_Fig8 | T0 | Visual Analogue Scale (VAS) | Craving | -1.15 |
| **(Li, Hartwell, et al., 2013)** | **(10)** | Nicotine | 14 | 14 | 6 cm rule | 100 | 1 | rTMS | 10 | 3000 | Magstim_70mm_Fig8 | T0 | Visual Analogue Scale (VAS) | Craving | 0.31 |
| **(Dieler et al., 2014)** | **(52)** | Nicotine | 36 | 38 | F4 | 80 | 4 | iTBS | 5 | 600 | MagVenture_MC-B70 | T0 | Questionnaire of Smoking Urges (QSU) | Craving | -0.19 |
| **(Ceccanti et al., 2015)** | **(31)** | Alcohol | 9 | 9 | F1 | 120 | 10 | dTMS | 20 | 1500 | H1 | T0 | Visual Analogue Scale (VAS) | Craving | -0.04 |
| **(S. C. Herremans et al., 2015)** | **(53)** | Alcohol | 26 | 26 | 5 cm rule | 110 | 1 | rTMS | 20 | 1560 | Magstim_70mm_Fig8 | T0 | Ten-point Likert Scales (TLS) | Craving | 0.03 |
| **(Trojak et al., 2015)** | **(54)** | Nicotine | 18 | 18 | Neuronavigation | 120 | 10 | rTMS | 1 | 360 | MagVenture_MCF-B65 | T0 | Visual Analogue Scale (VAS) | Craving | -0.11 |
| **(Del Felice et al., 2016)** | **(1)** | Alcohol | 9 | 8 | F3 | 100 | 4 | rTMS | 10 | 1000 | Magstim_70mm_Fig8 | T0 | Visual Analogue Scale (VAS) | Craving | -0.04 |
| **(Shen et al., 2016)** | **(55)** | Opioids | 10 | 10 | Not Reported | 100 | 5 | rTMS | 10 | 2000 | Magstim_70mm_Fig8 | T0 | Visual Analogue Scale (VAS) | Craving | 2.49 |

| | | | | | | | | | | | | | | | |
|---|---|---|---|---|---|---|---|---|---|---|---|---|---|---|---|
| **(Hanlon et al., 2017)** | **(56)** | Cocaine | 25 | 25 | Fp1 | 110 | 1 | cTBS | 5 | 3600 | Magstim_70mm_Fig8 | T0 | Visual Analogue Scale (VAS) | Craving | 0.10 |
| **(Hanlon et al., 2017)** | **(56)** | Alcohol | 24 | 24 | Fp1 | 110 | 1 | cTBS | 5 | 3600 | Magstim_70mm_Fig8 | T0 | Visual Analogue Scale (VAS) | Craving | -0.08 |
| **(Sahlem et al., 2018)** | **(12)** | Cannabis | 16 | 16 | Beam F3 | 110 | 1 | rTMS | 10 | 4000 | MagVenture_Cool-B65 | T0 | Marijuana Craving Questionnaire (MCQ) | Craving | -0.27 |
| **(Su et al., 2017)** | **(57)** | Meth | 15 | 15 | 5 cm rule | 80 | 5 | rTMS | 10 | 1200 | MagMore_PMD45-EEG | T0 | Visual Analogue Scale (VAS) | Craving | 0.30 |
| **(Liang, Wang, & Yuan, 2018)** | **(58)** | Meth | 22 | 24 | Not Reported | 100 | 10 | rTMS | 10 | 2000 | MagVenture_MC-125_new | T0 | Visual Analogue Scale (VAS) | Craving | 4.49 |
| **(Jansen et al., 2019)** | **(59)** | Alcohol | 18 | 17 | Neuronavigation | 110 | 1 | rTMS | 10 | 3000 | Magstim_70mm_Fig8 | T0 | Alcohol Urge Questionnaire (AUQ) | Craving | 0.52 |
| **(Chen, Su, Li, et al., 2020)** | **(60)** | Meth | 19 | 18 | F3 | 110 | 10 | iTBS | 5 | 900 | MagVenture_Cool-B70 | T0 | Visual Analogue Scale (VAS) | Craving | 0.91 |
| **(Chen, Su, Li, et al., 2020)** | **(60)** | Meth | 19 | 18 | Fp1 | 100 | 10 | cTBS | 5 | 900 | MagVenture_Cool-B70 | T0 | Visual Analogue Scale (VAS) | Craving | 1.25 |
| **(Chen, Su, Li, et al., 2020)** | **(60)** | Meth | 19 | 19 | F3 + Fp1 | 110 + 100 | 10 | iTBS/ cTBS | 5 | 900 | MagVenture_Cool-B70 | T0 | Visual Analogue Scale (VAS) | Craving | 1.48 |
| **(Chen, Su, Jiang, et al., 2020)** | **(61)** | Meth | 10 | 15 | Beam F3 | 100% | 20 | iTBS | 5 | 900 | MagVenture_Cool-B70 | T0 | Visual Analogue Scale (VAS) | Craving | 0.76 |
| **(Liu et al., 2020)** | **(62)** | Opioids | 35 | 35 | F3 | 100 | 20 | rTMS | 10 | 2000 | MagMore_PMD45-EEG | T0 | Visual Analogue Scale (VAS) | Craving | 3.52 |
| **(Liu et al., 2020)** | **(62)** | Opioids | 35 | 29 | F3 | 100 | 20 | rTMS | 1 | 600 | MagMore_PMD45-EEG | T0 | Visual Analogue Scale (VAS) | Craving | 2.33 |
| **(Perini et al., 2020)** | **(35)** | Alcohol | 22 | 23 | AFz | 120 | 15 | dTMS | 10 | 1500 | H8 | T0 | Penn Alcohol Craving Scale (PACS) | Craving | 0.67 |
| **(Raikwar et al., 2020)** | **(63)** | Alcohol | 30 | 30 | Not Reported | 120 | 10 | rTMS | 10 | 800 | Magstim_70mm_Fig8 | T0 | Alcohol Craving Questionnaire (ACQ) | Craving | 0.29 |
| **(Su, Liu, et al., 2020)** | **(64)** | Meth | 30 | 30 | Beam F3 | 100 | 20 | iTBS | 2 | 900 | MagVenture_Cool-B70 | T0 | Visual Analogue Scale (VAS) | Craving | 0.81 |
| **(Su, Chen, Zhong, et al., 2020)** | **(2)** | Meth | 25 | 25 | F3 | 100 | 20 | iTBS | 5 | 900 | MagVenture_Cool-B70 | T0 | Visual Analogue Scale (VAS) | Craving | 0.81 |
| **(Su, Chen, Jiang, et al., 2020)** | **(65)** | Meth | 56 | 70 | F3 | 100 | 20 | iTBS | 5 | 900 | MagVenture_Cool-B70 | T0 | Visual Analogue Scale (VAS) | Craving | 1.20 |
| **(Yuan et al., 2020)** | **(66)** | Meth | 36 | 37 | 5 cm rule | 100 | 10 | rTMS | 1 | 600 | MagMore_PMD45-EEG | T0 | Visual Analogue Scale (VAS) | Craving | 0.61 |
| **(Abdelrahman et al., 2021)** | **(36)** | Nicotine | 20 | 20 | 7 cm rule | 80 | 10 | rTMS | 20 | 2000 | Magstim_70mm_Fig8 | T0 | Tobacco Craving Questionnaire (TCQ) | Craving | 1.10 |
| **(Chen et al., 2021)** | **(67)** | Meth | 19 | 30 | Not Reported | 100 | 20 | iTBS | 5 | 900 | MCF-B65(New) | T0 | Visual Analogue Scale (VAS) | Craving | 1.62 |
| **(Garza-Villarreal et al., 2021)** | **(68)** | Cocaine | 20 | 24 | Fiducial marker | 100 | 10 | rTMS | 5 | 5000 | MagVenture_Cool-B65 | T0 | Visual Analogue Scale (VAS) | Craving | 0.64 |
| **(Lolli et al., 2021)** | **(69)** | Cocaine | 27 | 30 | 5 cm rule | 100 | 15 | rTMS | 15 | 2400 | MagVenture_Cool-B65 | T0 | Visual Analogue Scale (VAS) | Craving | -0.03 |
| **(Tsai et al., 2021)** | **(70)** | Opioids | 9 | 11 | 5 cm rule | 100 | 11 | rTMS | 15 | 2400 | Magstim_70mm_Fig8 | T0 | Visual Analogue Scale (VAS) | Craving | -0.31 |

| | | | | | | | | | | | | | | |
|---|---|---|---|---|---|---|---|---|---|---|---|---|---|---|
| **(Zangen et al., 2021)** | **(71)** | Nicotine | 139 | 123 | Fz | 120 | 15 | dTMS | 10 | 1800 | H4 | T0 | Visual Analogue Scale (VAS) | Craving | 3.92 |
| **(Ankit et al., 2022)** | **(72)** | Opioids | 20 | 20 | Fp2 | 80 | 14 | cTBS | 5 | 900 | Cool-DB80 | T0 | Obsessive Compulsive Drug Use Scale (OCDUS) | Craving | 0.22 |
| **(Belgers et al., 2022)** | **(37)** | Alcohol | 16 | 14 | F4 | 110 | 10 | rTMS | 10 | 3000 | Magstim_70mm_Fig8 | T0 | Visual Analogue Scale (VAS) | Craving | 0.65 |
| **(Bidzinski et al., 2022)** | **(4)** | Cannabis | 8 | 9 | F5 + F6 | 90 | 20 | rTMS | 20 | 750 | MagVenture_Cool-B65 | T0 | Marijuana Craving Questionnaire (MCQ) | Craving | 0.42 |
| **(Harel et al., 2022)** | **(73)** | Alcohol | 23 | 23 | AFz | 100 | 15 | dTMS | 10 | 3000 | H7 | T0 | Penn Alcohol Craving Scale (PACS) | Craving | 0.23 |
| **(Jin et al., 2022)** | **(8)** | Opioids | 18 | 17 | 6 cm rule | 100 | 7 | rTMS | 10 | 2000 | Magstim_70mm_Fig8 | T0 | Visual Analogue Scale (VAS) | Craving | 0.53 |
| **(Marques et al., 2022)** | **(74)** | Nicotine | 11 | 12 | Fp1 | 110 | 1 | rTMS | 1 | 1200 | MagVenture_Cool-B70 | T0 | Visual Analogue Scale (VAS) | Craving | 0.53 |
| **(Marques et al., 2022)** | **(74)** | Nicotine | 11 | 12 | Cz | 90 | 1 | rTMS | 1 | 1200 | MagVenture_Cool-B70 | T0 | Visual Analogue Scale (VAS) | Craving | 0.53 |
| **(McNeill et al., 2022)** | **(3)** | Alcohol | 20 | 20 | F3 | 80 | 1 | cTBS | 5 | 600 | Magstim_70mm_Fig8 | T0 | Desire for Alcohol Questionnaire (DAQ) | Craving | -1.06 |
| **(McNeill et al., 2022)** | **(3)** | Alcohol | 20 | 20 | F4 | 80 | 1 | cTBS | 5 | 600 | Magstim_70mm_Fig8 | T0 | Desire for Alcohol Questionnaire (DAQ) | Craving | -0.30 |
| **(McNeill et al., 2022)** | **(3)** | Alcohol | 20 | 20 | Fpz | 80 | 1 | cTBS | 5 | 600 | Magstim_70mm_Fig8 | T0 | Desire for Alcohol Questionnaire (DAQ) | Craving | -0.24 |
| **(Mikellides et al., 2022)** | **(38)** | Nicotine | 30 | 30 | Beam F3 | 100 | 20 | iTBS | 5 | 600 | MagVenture_Cool-B65 | T0 | Visual Analogue Scale (VAS) | Craving | -0.02 |
| **(Li et al., 2022)** | **(40)** | Nicotine | 4 | 3 | 5.5 cm rule | 100 | 5 | rTMS | 10 | 3000 | Magstim_70mm_Fig8 | T0 | Questionnaire of Smoking Urges (QSU) | Craving | 0.34 |
| **(Kang et al., 2022)** | **(7)** | Opioids | 20 | 22 | 5 cm rule | 80 | 30 | iTBS | 5 | 600 | MagMore_PMD70 | T0 | Visual Analogue Scale (VAS) | Craving | 0.82 |
| **(Wang et al., 2022)** | **(75)** | Meth | 30 | 34 | F4 | 100 | 12 | rTMS | 10 | 400 | Magstim_70mm_Fig8 | T0 | Visual Analogue Scale (VAS) | Craving | 0.51 |
| **(McCalley et al., 2025)** | **(76)** | Alcohol | 20 | 21 | Fp1 | 110 | 10 | cTBS | 5 | 3600 | MagVenture_Cool-B65 | T0 | Obsessive Compulsive Drinking Scale (OCDS) | Craving | 0.61 |
| **(Hoven et al., 2023)** | **(77)** | Alcohol | 38 | 33 | F4 | 110 | 10 | rTMS | 10 | 3000 | Magstim_70mm_Fig8 | T0 | Obsessive Compulsive Drinking Scale (OCDS) | Craving | 0.00 |
| **(Ibrahim et al., 2023)** | **(41)** | Nicotine | 18 | 24 | NA | 120 | 20 | dTMS | 10 | 1020 | H11 | T0 | Questionnaire of Smoking Urges (QSU) | Craving | -0.07 |
| **(Gong et al., 2023)** | **(78)** | Opioids | 38 | 38 | NA | 100 | 20 | iTBS | 20 | 900 | Not Reported | T0 | Visual Analogue Scale (VAS) | Craving | 0.73 |
| **(Addicott et al., 2024)** | **(45)** | Nicotine | 13 | 25 | AFz | 110% | 20+8 | iTBS | 5 | 600 | MagVenture_Cool-B65 | T0 | Questionnaire of Smoking Urges (QSU) | Craving | 2.45 |
| **(Bellini et al., 2024)** | **(44)** | Nicotine | 37 | 41 | Fz | 120 | 21 | dTMS | 10 | 1800 | H4 | T0 | Tobacco Craving Questionnaire (TCQ) | Craving | 0.19 |
| **(S. Li, X. Ma, et al., 2024)** | **(79)** | Nicotine | 18 | 43 | 5 cm rule | 100 | 5 | rTMS | 10 | 3000 | Magstim_70mm_Fig8 | T0 | Tiffany Questionnaire on Smoking Urges (TQSU) | Craving | 1.68 |

| | | | | | | | | | | | | | | | |
|---|---|---|---|---|---|---|---|---|---|---|---|---|---|---|---|
| **(Padula et al., 2024)** | **(80)** | Alcohol | 9 | 8 | Beam F3 | 110 | 20 | iTBS | 5 | 600 | MagVenture_Cool-B65 | T0 | Obsessive Compulsive Drug Use Scale (OCDUS) | Craving | -2.99 |
| **(Rakesh et al., 2024)** | **(81)** | Nicotine | 5 | 8 | Neuronavigation | 120 | 1 | iTBS | 5 | 1800 | MagVenture_Cool-B65 | T0 | Tobacco Craving Questionnaire (TCQ) | Craving | 0.47 |
| **(Rasgado-Toledo et al., 2024)** | **(82)** | Cocaine | 20 | 25 | 5.5 cm rule | 100 | 20 | rTMS | 5 | 5000 | MagVenture_Cool-B65 | T0 | Visual Analogue Scale (VAS) | Craving | 0.49 |
| **(S. Li, Z. Zhang, et al., 2024)** | **(83)** | Nicotine | 18 | 42 | 5 cm rule | 100 | 5 | rTMS | 10 | 2000 | Magstim_70mm_Fig8 | T0 | Tiffany Questionnaire on Smoking Urges (TQSU) | Craving | 1.32 |
| **(Sahlem et al., 2024)** | **(11)** | Cannabis | 23 | 28 | Beam F3 | 120 | 20 | rTMS | 10 | 4000 | MagVenture_Cool-B65 | T0 | Marijuana Craving Questionnaire (MCQ) | Craving | 0.26 |
| **(Y. Li et al., 2024)** | **(84)** | Meth | 17 | 17 | 5 cm rule | 100 | 8 | rTMS | 10 | 1200 | Magstim_70mm_Fig8 | T0 | Visual Analogue Scale (VAS) | Craving | 1.02 |
| **(Ward et al., 2025)** | **(13)** | Nicotine | 10 | 10 | fMRI | 100 | 1 | iTBS | 5 | 600 | MagVenture_Cool-B65 | T0 | Visual Analogue Scale (VAS) | Craving | 2.10 |
| **(Ward et al., 2025)** | **(13)** | Nicotine | 10 | 10 | fMRI | 80 | 1 | cTBS | 5 | 600 | MagVenture_Cool-B65 | T0 | Visual Analogue Scale (VAS) | Craving | 0.00 |
| **(Belgers et al., 2025)** | **(85)** | Alcohol | 16 | 14 | F4 | 110 | 10 | rTMS | 10 | 3000 | Magstim_70mm_Fig8 | T0 | Visual Analogue Scale (VAS) | Craving | 0.65 |
| **(Tang et al., 2025)** | **(86)** | Opioids | 2 | 2 | Neuronavigation | 120 | 20 | iTBS/ cTBS | 5 | 600 | MagVenture_Cool-B70 | T0 | Visual Analogue Scale (VAS) | Craving | 0.83 |
| **(D. M. McCalley et al., 2025)** | **(47)** | Cocaine | 16 | 17 | Fp1 | 110 | 10 | cTBS | 5 | 1800 | MagVenture_Cool-B65 | T0 | Cocaine Craving Questionnaire (CCQ) | Craving | 0.17 |
| **(Ghazi, et al., 2025)** | **(87)** | Cocaine | 20 | 24 | Beam F3 | 110 | 10 | rTMS | 10 | 3000 | MagVenture_Cool-B65 | T0 | Visual Analogue Scale (VAS) | Craving | 1.50 |
| **(Petersen et al., 2025)** | **(14)** | Nicotine | 60 | 66 | fMRI | 100 | 1 | rTMS | 10 | 3000 | MagVenture_Cool-B65 | T0 | Urge to Smoke Scale (UTS) | Craving | 0.29 |
| **(Petersen et al., 2025)** | **(14)** | Nicotine | 61 | 66 | fMRI | 100 | 1 | rTMS | 10 | 3000 | MagVenture_Cool-B65 | T0 | Urge to Smoke Scale (UTS) | Craving | 0.32 |
| **(Petersen et al., 2025)** | **(14)** | Nicotine | 62 | 66 | fMRI | 100 | 1 | rTMS | 10 | 3000 | MagVenture_Cool-B65 | T0 | Urge to Smoke Scale (UTS) | Craving | 0.10 |
| **(Guldas et al., 2025)** | **(48)** | Opioids | 18 | 21 | 5 cm rule | 110 | 20 | rTMS | 10 | 3000 | Cool-DB80 | T0 | Visual Analogue Scale (VAS) | Craving | 0.36 |

**Reference**


1. Del Felice A, Bellamoli E, Formaggio E, Manganotti P, Masiero S, Cuoghi G, et al. Neurophysiological, psychological and behavioural correlates of rTMS treatment in alcohol dependence. Drug and alcohol dependence. 2016;158:147-53.
2. Su H, Chen T, Zhong N, Jiang H, Du J, Xiao K, et al. γ-aminobutyric acid and glutamate/glutamine alterations of the left prefrontal cortex in individuals with methamphetamine use disorder: a combined transcranial magnetic stimulation-magnetic resonance spectroscopy study. Annals of translational medicine. 2020;8(6):347.
3. McNeill AM, Monk RL, Qureshi AW, Makris S, Cazzato V, Heim D. Elevated ad libitum alcohol consumption following continuous theta burst stimulation to the left-dorsolateral prefrontal cortex is partially mediated by changes in craving. Cognitive, Affective, & Behavioral Neuroscience. 2022;22(1):160-70.
4. Bidzinski KK, Lowe DJ, Sanches M, Sorkhou M, Boileau I, Kiang M, et al. Investigating repetitive transcranial magnetic stimulation on cannabis use and cognition in people with schizophrenia. Schizophrenia. 2022;8(1):2.
5. Eichhammer P, Johann M, Kharraz A, Binder H, Pittrow D, Wodarz N, et al. High-frequency repetitive transcranial magnetic stimulation decreases cigarette smoking. Journal of Clinical Psychiatry. 2003;64(8):951-3.
6. Amiaz R, Levy D, Vainiger D, Grunhaus L, Zangen A. Repeated high-frequency transcranial magnetic stimulation over the dorsolateral prefrontal cortex reduces cigarette craving and consumption. Addiction. 2009;104(4):653-60.
7. Kang T, Ding X, Zhao J, Li X, Xie R, Jiang H, et al. Influence of improved behavioral inhibition on decreased cue-induced craving in heroin use disorder: a preliminary intermittent theta burst stimulation study. Journal of Psychiatric Research. 2022;152:375-83.
8. Jin L, Yuan M, Zhang W, Su H, Wang F, Zhu J, et al. Repetitive transcranial magnetic stimulation modulates coupling among large-scale brain networks in heroin-dependent individuals: A randomized resting-state functional magnetic resonance imaging study. Addiction Biology. 2022;27(2):e13121.
9. Li X, Malcolm RJ, Huebner K, Hanlon CA, Taylor JJ, Brady KT, et al. Low frequency repetitive transcranial magnetic stimulation of the left dorsolateral prefrontal cortex transiently increases cue-induced craving for methamphetamine: a preliminary study. Drug and alcohol dependence. 2013;133(2):641-6.
10. Li X, Hartwell KJ, Owens M, LeMatty T, Borckardt JJ, Hanlon CA, et al. Repetitive transcranial magnetic stimulation of the dorsolateral prefrontal cortex reduces nicotine cue craving. Biological psychiatry. 2013;73(8):714-20.
11. Sahlem GL, Kim B, Baker NL, Wong BL, Caruso MA, Campbell LA, et al. A preliminary randomized controlled trial of repetitive transcranial magnetic stimulation applied to the left dorsolateral prefrontal cortex in treatment seeking participants with cannabis use disorder. Drug and alcohol dependence. 2024;254:111035.
12. Sahlem GL, Baker NL, George MS, Malcolm RJ, McRae-Clark AL. Repetitive transcranial magnetic stimulation (rTMS) administration to heavy cannabis users. The American journal of drug and alcohol abuse. 2018;44(1):47-55.

13. Ward HB, Blyth SH, Vandekar S, Rogers BP, Yildiz G, Connolly JG, et al. Defining and Engaging a Novel rTMS Target for Nicotine Craving in Psychotic Disorders. medRxiv. 2025:2025.09. 24.25334160.
14. Petersen N, Apostol MR, Jordan T, Ngo TDP, Kearley NW, London ED, et al. Comparing neuromodulation targets to reduce cigarette craving and withdrawal: a randomized clinical trial. Neuropsychopharmacology. 2025:1-8.
15. Li X, Hartwell KJ, Henderson S, Badran BW, Brady KT, George MS. Two weeks of image-guided left dorsolateral prefrontal cortex repetitive transcranial magnetic stimulation improves smoking cessation: A double-blind, sham-controlled, randomized clinical trial. Brain stimulation. 2020;13(5):1271-9.
16. Herremans S, Baeken C, Vanderbruggen N, Vanderhasselt M-A, Zeeuws D, Santermans L, et al. No influence of one right-sided prefrontal HF-rTMS session on alcohol craving in recently detoxified alcohol-dependent patients: results of a naturalistic study. Drug and alcohol dependence. 2012;120(1-3):209-13.
17. Caulfield KA, Fleischmann HH, Cox CE, Wolf JP, George MS, McTeague LM. Neuronavigation maximizes accuracy and precision in TMS positioning: Evidence from 11,230 distance, angle, and electric field modeling measurements. Brain stimulation. 2022;15(5):1192-205.
18. Soleimani G, Souki A, Honari S, Baker TE, Brunoni AR, Ebrahimi M, et al. Effectiveness of Noninvasive Brain Stimulation Protocols on Drug Craving and Consumption/Relapse in Substance Use Disorders: A Systematic Review and Meta-analysis of 208 Clinical Trials and 36 Protocols. medRxiv. 2025:2025.09. 21.25335559.
19. Saturnino GB, Thielscher A, Madsen KH, Knösche TR, Weise K. A principled approach to conductivity uncertainty analysis in electric field calculations. Neuroimage. 2019;188:821-34.
20. McCann H, Pisano G, Beltrachini L. Variation in reported human head tissue electrical conductivity values. Brain topography. 2019;32:825-58.
21. Wischnewski M, Berger TA, Opitz A, Alekseichuk I. Causal functional maps of brain rhythms in working memory. Proceedings of the National Academy of Sciences. 2024;121(14):e2318528121.
22. Ekhtiari H, Kuplicki R, Pruthi A, Paulus M. Methamphetamine and Opioid Cue Database (MOCD): Development and Validation. Drug and alcohol dependence. 2020:107941.
23. Baetens K, Van Hoornweder S, Berger TA, Wischnewski M. ACES: Automated Correlation of Electric field strength and Stimulation effects for non-invasive brain stimulation. Brain Stimulation: Basic, Translational, and Clinical Research in Neuromodulation. 2024;17(2):473-5.
24. Wischnewski M, Mantell KE, Opitz A. Identifying regions in prefrontal cortex related to working memory improvement: A novel meta-analytic method using electric field modeling. Neuroscience & Biobehavioral Reviews. 2021;130:147-61.
25. Wischnewski M, Berger TA, Opitz A. Meta-modeling the effects of anodal left prefrontal transcranial direct current stimulation on working memory performance. Imaging Neuroscience. 2024;2:1-14.
26. Sinanaj L, Pallis K, Dehkordi AF, Huguelet P, Kaiser S, Bègue I. Mapping symptom-general and symptom-specific targets for transcranial magnetic stimulation in schizophrenia: an electric-field modeling meta-analysis. Molecular Psychiatry. 2025:1-11.

27. Kuhn T, Indahlastari A, Vila-Rodriguez F, Fonzo G, Petersen N, Rotstein N, et al. The ENIGMA-Neuromodulation working group–A mission statement. Brain Stimulation: Basic, Translational, and Clinical Research in Neuromodulation. 2025;18(2):142-4.
28. Sheffer CE, Mennemeier M, Landes RD, Bickel WK, Brackman S, Dornhoffer J, et al. Neuromodulation of delay discounting, the reflection effect, and cigarette consumption. Journal of substance abuse treatment. 2013;45(2):206-14.
29. Dinur-Klein L, Dannon P, Hadar A, Rosenberg O, Roth Y, Kotler M, et al. Smoking cessation induced by deep repetitive transcranial magnetic stimulation of the prefrontal and insular cortices: a prospective, randomized controlled trial. Biological psychiatry. 2014;76(9):742-9.
30. Prikryl R, Ustohal L, Kucerova HP, Kasparek T, Jarkovsky J, Hublova V, et al. Repetitive transcranial magnetic stimulation reduces cigarette consumption in schizophrenia patients. Progress in Neuro-Psychopharmacology and Biological Psychiatry. 2014;49:30-5.
31. Ceccanti M, Inghilleri M, Attilia ML, Raccah R, Fiore M, Zangen A, et al. Deep TMS on alcoholics: effects on cortisolemia and dopamine pathway modulation. A pilot study. Canadian journal of physiology and pharmacology. 2015;93(4):283-90.
32. Bolloni C, Panella R, Pedetti M, Frascella AG, Gambelunghe C, Piccoli T, et al. Bilateral transcranial magnetic stimulation of the prefrontal cortex reduces cocaine intake: a pilot study. Frontiers in psychiatry. 2016;7:133.
33. Huang W, Fang S, Zhang J, Baoping X. Effect of repetitive transcranial magnetic stimulation on cigarette smoking in patients with schizophrenia. Shanghai archives of psychiatry. 2016;28(6):309.
34. Addolorato G, Antonelli M, Cocciolillo F, Vassallo GA, Tarli C, Sestito L, et al. Deep transcranial magnetic stimulation of the dorsolateral prefrontal cortex in alcohol use disorder patients: effects on dopamine transporter availability and alcohol intake. European Neuropsychopharmacology. 2017;27(5):450-61.
35. Perini I, Kämpe R, Arlestig T, Karlsson H, Löfberg A, Pietrzak M, et al. Repetitive transcranial magnetic stimulation targeting the insular cortex for reduction of heavy drinking in treatment-seeking alcohol-dependent subjects: a randomized controlled trial. Neuropsychopharmacology. 2020;45(5):842-50.
36. Abdelrahman AA, Noaman M, Fawzy M, Moheb A, Karim AA, Khedr EM. A double-blind randomized clinical trial of high frequency rTMS over the DLPFC on nicotine dependence, anxiety and depression. Scientific Reports. 2021;11(1):1640.
37. Belgers M, Van Eijndhoven P, Markus W, Schene AH, Schellekens A. rTMS reduces craving and alcohol use in patients with alcohol use disorder: results of a randomized, sham-controlled clinical trial. Journal of clinical medicine. 2022;11(4):951.
38. Mikellides G, Michael P, Psalta L, Stefani A, Schuhmann T, Sack AT. Accelerated intermittent theta burst stimulation in smoking cessation: Placebo effects equal to active stimulation when using advanced placebo coil technology. Frontiers in psychiatry. 2022;13:892075.
39. Moeller SJ, Gil R, Weinstein JJ, Baumvoll T, Wengler K, Fallon N, et al. Deep rTMS of the insula and prefrontal cortex in smokers with schizophrenia: proof-of-concept study. Schizophrenia. 2022;8(1):6.

40. Li X, Toll BA, Carpenter MJ, Nietert PJ, Dancy M, George MS, et al. Repetitive transcranial magnetic stimulation for tobacco treatment in cancer patients: a preliminary report of a one-week treatment. Journal of Smoking Cessation. 2022;2022:e9.
41. Ibrahim C, Tang VM, Blumberger DM, Malik S, Tyndale RF, Trevizol AP, et al. Efficacy of insula deep repetitive transcranial magnetic stimulation combined with varenicline for smoking cessation: a randomized, double-blind, sham controlled trial. Brain Stimulation. 2023;16(5):1501-9.
42. Li X, Caulfield KA, Hartwell KJ, Henderson S, Brady KT, George MS. Reduced executive and reward connectivity is associated with smoking cessation response to repetitive transcranial magnetic stimulation: A double-blind, randomized, sham-controlled trial. Brain imaging and behavior. 2024;18(1):207-19.
43. Du X, Choa F-S, Chiappelli J, Bruce H, Kvarta M, Summerfelt A, et al. Combining neuroimaging and brain stimulation to test alternative causal pathways for nicotine addiction in schizophrenia. Brain stimulation. 2024;17(2):324-32.
44. Bellini BB, Scholz JR, Abe TO, Arnaut D, Tonstad S, Alberto RL, et al. Does deep TMS really works for smoking cessation? A prospective, double blind, randomized, sham controlled study. Progress in Neuro-Psychopharmacology and Biological Psychiatry. 2024;132:110997.
45. Addicott MA, Kinney KR, Saldana S, Ip EH-S, DeMaioNewton H, Bickel WK, et al. A randomized controlled trial of intermittent theta burst stimulation to the medial prefrontal cortex for tobacco use disorder: clinical efficacy and safety. Drug and alcohol dependence. 2024;258:111278.
46. Durazzo TC, Kraybill EP, Stephens LH, McCalley DM, Humphreys K, May AC, et al. Intermittent theta burst to the left dorsolateral prefrontal cortex promoted decreased alcohol consumption and improved outcomes in those with alcohol use disorder: A randomized, double-blind, placebo-controlled clinical trial. Drug and alcohol dependence. 2025;270:112641.
47. McCalley DM, Kinney KR, Kaur N, Wolf JP, Contreras IE, Smith JP, et al. A randomized controlled trial of medial prefrontal cortex theta burst stimulation for cocaine use disorder: a three-month feasibility and brain target engagement study. Biological Psychiatry: Cognitive Neuroscience and Neuroimaging. 2025;10(6):616-25.
48. Guldas S, Tumkaya S, Yucens B. Deep Transcranial Magnetic Stimulation in Patients With Opioid Use Disorder: A Double-Blind, Placebo-Controlled Randomized Trial. Addiction Biology. 2025;30(6):e70057.
49. Mishra BR, Nizamie SH, Das B, Praharaj SK. Efficacy of repetitive transcranial magnetic stimulation in alcohol dependence: a sham-controlled study. Addiction. 2010;105(1):49-55.
50. Rose JE, McClernon FJ, Froeliger B, Behm FM, Preud'homme X, Krystal AD. Repetitive transcranial magnetic stimulation of the superior frontal gyrus modulates craving for cigarettes. Biological psychiatry. 2011;70(8):794-9.
51. Herremans S, Vanderhasselt M-A, De Raedt R, Baeken C. Reduced intra-individual reaction time variability during a go–NoGo task in detoxified alcohol-dependent patients after one right-sided dorsolateral prefrontal HF-rTMS session. Alcohol and alcoholism. 2013;48(5):552-7.
52. Dieler AC, Dresler T, Joachim K, Deckert J, Herrmann MJ, Fallgatter AJ. Can intermittent theta burst stimulation as add-on to psychotherapy improve nicotine abstinence? Results from a pilot study. European addiction research. 2014;20(5):248-53.

53. Herremans SC, Van Schuerbeek P, De Raedt R, Matthys F, Buyl R, De Mey J, et al. The impact of accelerated right prefrontal high-frequency repetitive transcranial magnetic stimulation (rTMS) on cue-reactivity: an fMRI study on craving in recently detoxified alcohol-dependent patients. PLoS One. 2015;10(8):e0136182.
54. Trojak B, Meille V, Achab S, Lalanne L, Poquet H, Ponavoy E, et al. Transcranial magnetic stimulation combined with nicotine replacement therapy for smoking cessation: a randomized controlled trial. Brain stimulation. 2015;8(6):1168-74.
55. Shen Y, Cao X, Tan T, Shan C, Wang Y, Pan J, et al. 10-Hz repetitive transcranial magnetic stimulation of the left dorsolateral prefrontal cortex reduces heroin cue craving in long-term addicts. Biological psychiatry. 2016;80(3):e13-e4.
56. Hanlon CA, Dowdle LT, Correia B, Mithoefer O, Kearney-Ramos T, Lench D, et al. Left frontal pole theta burst stimulation decreases orbitofrontal and insula activity in cocaine users and alcohol users. Drug and alcohol dependence. 2017;178:310-7.
57. Su H, Zhong N, Gan H, Wang J, Han H, Chen T, et al. High frequency repetitive transcranial magnetic stimulation of the left dorsolateral prefrontal cortex for methamphetamine use disorders: a randomised clinical trial. Drug and alcohol dependence. 2017;175:84-91.
58. Liang Y, Wang L, Yuan T-F. Targeting withdrawal symptoms in men addicted to methamphetamine with transcranial magnetic stimulation: a randomized clinical trial. JAMA psychiatry. 2018;75(11):1199-201.
59. Jansen JM, Van den Heuvel OA, Van der Werf YD, De Wit SJ, Veltman DJ, Van den Brink W, et al. The effect of high-frequency repetitive transcranial magnetic stimulation on emotion processing, reappraisal, and craving in alcohol use disorder patients and healthy controls: a functional magnetic resonance imaging study. Frontiers in Psychiatry. 2019;10:272.
60. Chen T, Su H, Li R, Jiang H, Li X, Wu Q, et al. The exploration of optimized protocol for repetitive transcranial magnetic stimulation in the treatment of methamphetamine use disorder: a randomized sham-controlled study. EBioMedicine. 2020;60.
61. Chen T, Su H, Jiang H, Li X, Zhong N, Du J, et al. Cognitive and emotional predictors of real versus sham repetitive transcranial magnetic stimulation treatment response in methamphetamine use disorder. Journal of psychiatric research. 2020;126:73-80.
62. Liu X, Zhao X, Liu T, Liu Q, Tang L, Zhang H, et al. The effects of repetitive transcranial magnetic stimulation on cue-induced craving in male patients with heroin use disorder. EBioMedicine. 2020;56.
63. Raikwar S, Divinakumar K, Prakash J, Khan SA, GuruPrakash K, Batham S. A sham-controlled trial of repetitive transcranial magnetic stimulation over left dorsolateral prefrontal cortex and its effects on craving in patients with alcohol dependence. Industrial Psychiatry Journal. 2020;29(2):245-50.
64. Su H, Liu Y, Yin D, Chen T, Li X, Zhong N, et al. Neuroplastic changes in resting-state functional connectivity after rTMS intervention for methamphetamine craving. Neuropharmacology. 2020;175:108177.
65. Su H, Chen T, Jiang H, Zhong N, Du J, Xiao K, et al. Intermittent theta burst transcranial magnetic stimulation for methamphetamine addiction: a randomized clinical trial. European Neuropsychopharmacology. 2020;31:158-61.

66. Yuan J, Liu W, Liang Q, Cao X, Lucas MV, Yuan T-F. Effect of low-frequency repetitive transcranial magnetic stimulation on impulse inhibition in abstinent patients with methamphetamine addiction: a randomized clinical trial. JAMA network open. 2020;3(3):e200910-e.
67. Chen T, Su H, Wang L, Li X, Wu Q, Zhong N, et al. Modulation of methamphetamine-related attention bias by intermittent theta-burst stimulation on left dorsolateral prefrontal cortex. Frontiers in Cell and Developmental Biology. 2021;9:667476.
68. Garza-Villarreal EA, Alcala-Lozano R, Fernandez-Lozano S, Morelos-Santana E, Dávalos A, Villicaña V, et al. Clinical and functional connectivity outcomes of 5-Hz repetitive transcranial magnetic stimulation as an add-on treatment in cocaine use disorder: a double-blind randomized controlled trial. Biological Psychiatry: Cognitive Neuroscience and Neuroimaging. 2021;6(7):745-57.
69. Lolli F, Salimova M, Scarpino M, Lanzo G, Cossu C, Bastianelli M, et al. A randomised, double-blind, sham-controlled study of left prefrontal cortex 15 Hz repetitive transcranial magnetic stimulation in cocaine consumption and craving. PLoS One. 2021;16(11):e0259860.
70. Tsai T-Y, Wang T-Y, Liu YC, Lee P-W, Chang WH, Lu T-H, et al. Add-on repetitive transcranial magnetic stimulation in patients with opioid use disorder undergoing methadone maintenance therapy. The American Journal of Drug and Alcohol Abuse. 2021;47(3):330-43.
71. Zangen A, Moshe H, Martinez D, Barnea-Ygael N, Vapnik T, Bystritsky A, et al. Repetitive transcranial magnetic stimulation for smoking cessation: a pivotal multicenter double-blind randomized controlled trial. World Psychiatry. 2021;20(3):397-404.
72. Ankit A, Das B, Dey P, Kshitiz KK, Khess CRJ. Efficacy of continuous theta burst stimulation-repetitive trancranial magnetic stimulation on the orbito frontal cortex as an adjunct to naltrexone in patients of opioid use disorder and its correlation with serum BDNF levels: a sham-controlled study. Journal of addictive diseases. 2022;40(3):373-81.
73. Harel M, Perini I, Kämpe R, Alyagon U, Shalev H, Besser I, et al. Repetitive transcranial magnetic stimulation in alcohol dependence: a randomized, double-blind, sham-controlled proof-of-concept trial targeting the medial prefrontal and anterior cingulate cortices. Biological psychiatry. 2022;91(12):1061-9.
74. Marques RC, Marques D, Vieira L, Cantilino A. Left frontal pole repetitive transcranial magnetic stimulation reduces cigarette cue-reactivity in correlation with verbal memory performance. Drug and Alcohol Dependence. 2022;235:109450.
75. Wang W, Zhu Y, Wang L, Mu L, Zhu L, Ding D, et al. High-frequency repetitive transcranial magnetic stimulation of the left dorsolateral prefrontal cortex reduces drug craving and improves decision-making ability in methamphetamine use disorder. Psychiatry research. 2022;317:114904.
76. McCalley D, Kinney K, Hanlon C. Medial prefrontal cortex theta burst stimulation improves treatment outcomes in cocaine dependence: a double-blind, sham-controlled neuroimaging study. Brain Stimulation: Basic, Translational, and Clinical Research in Neuromodulation. 2025;18(1):358.
77. Hoven M, Schluter RS, Schellekens AF, van Holst RJ, Goudriaan AE. Effects of 10 add-on HF-rTMS treatment sessions on alcohol use and craving among detoxified inpatients with alcohol use disorder: a randomized sham-controlled clinical trial. Addiction. 2023;118(1):71-85.

78. Gong H, Huang Y, Zhu X, Lu W, Cai Z, Zhu N, et al. Impact of combination of intermittent Theta burst stimulation and methadone maintenance treatment in individuals with opioid use disorder: a comparative study. Psychiatry Research. 2023;327:115411.
79. Li S, Ma X, Chen Ha, Wang M, Zheng Y, Yang B, et al. rTMS effects on urges and severity of tobacco use disorder operate independently of a retrieval-extinction component and involve frontal-striatal pathways. Journal of Affective Disorders. 2024;349:21-31.
80. Padula CB, McCalley DM, Tenekedjieva LT, MacNiven K, Rauch A, Morales JM, et al. A pilot, randomized clinical trial: left dorsolateral prefrontal cortex intermittent theta burst stimulation improves treatment outcomes in veterans with alcohol use disorder. Alcohol: Clinical and Experimental Research. 2024;48(1):164-77.
81. Rakesh G, Adams TG, Morey RA, Alcorn III JL, Khanal R, Su AE, et al. Intermittent theta burst stimulation and functional connectivity in people living with HIV/AIDS who smoke tobacco cigarettes: a preliminary pilot study. Frontiers in psychiatry. 2024;15:1315854.
82. Rasgado-Toledo J, Issa-Garcia V, Alcalá-Lozano R, Garza-Villarreal EA, González-Escamilla G. Cortical and subcortical microstructure integrity changes after repetitive transcranial magnetic stimulation therapy in cocaine use disorder and relates to clinical outcomes. Addiction Biology. 2024;29(2):e13381.
83. Li S, Zhang Z, Jiang A, Ma X, Wang M, Ni H, et al. Repetitive transcranial magnetic stimulation reshaped the dynamic reconfiguration of the executive and reward networks in individuals with tobacco use disorder. Journal of Affective Disorders. 2024;365:427-36.
84. Li Y, Yang B, Ma J, Li Y, Zeng H, Zhang J. Assessment of rTMS treatment effects for methamphetamine addiction based on EEG functional connectivity. Cognitive Neurodynamics. 2024;18(5):2373-86.
85. Belgers M, Markus W, Grasso F, Arns M, Van Eijndhoven P, Schellekens A. No Effects of rTMS on Performance Monitoring and Attentional Bias in Patients With Alcohol Use Disorder: A Pilot Study. Addiction Biology. 2025;30(11):e70100.
86. Tang VM, Le Foll B, Daskalakis ZJ, Wang A-L, Buckley L, Blumberger DM, et al. Repetitive Transcranial Magnetic Stimulation for the Treatment of Suicidality in Opioid Use Disorder: A Pilot Feasibility Randomized Controlled Trial. European Psychiatry. 2025:1-26.
87. Ghazi N, Garza-Villarreal EA, Soltanian-Zadeh H. Brain connectomics markers for response prediction to transcranial magnetic stimulation in cocaine use disorder. Scientific Reports. 2025;15(1):15336.
